# Genome-wide association studies reveal differences in genetic susceptibility between single events versus recurrent events of atrial fibrillation and myocardial infarction: the HUNT study

**DOI:** 10.1101/2024.01.17.24301410

**Authors:** Martina Hall, Anne Heidi Skogholt, Ida Surakka, Haavard Dalen, Eivind Almaas

**Affiliations:** Department of Biotechnology and Food Science, Norwegian University of Science and Technology, Trondheim, Norway; K. G. Jebsen center for Genetic Epidemiology, Norwegian University of Science and Technology, Trondheim, Norway; Department of Internal Medicine, University of Michigan, Ann Arbor, Michigan, USA; Clinic of Cardiology, St. Olavs University Hospital, Trondheim, Norway; Department of Circulation and Medical Imaging, Norwegian University of Science and Technology, Trondheim, Norway; Department of Medicine, Levanger Hospital, Nord-Trøndelag Hospital Trust, Levanger, Norway

**Keywords:** GWAS meta-analysis, recurrent events, atrial fibrillation, myocardial infarction, networks, PheWAS, HUNT

## Abstract

Genetic research into Atrial Fibrillation (AF) and Myocardial Infarction (MI) has predominantly concentrated on contrasting afflicted individuals with their healthy counterparts. However, this approach lacks granularity, overlooking the subtleties within patient populations. In this study, we explore the distinction between AF and MI patients experiencing only single events to those experiencing recurrent events. Integrating hospital records, questionnaire data, clinical measurements, and genetic data from more than 500, 000 HUNT and UKBB participants in our analysis, we compare the two groups for both clinical and genetic characteristics with GWAS meta-analyses, PheWAS-analyses, and gene co-expression networks. We find that the two groups of patients differs in both clinical characteristics and genetic risk. More specifically, recurrent AF patients are significantly younger with a better baseline health, in terms of lower cholesterol and blood pressure, than the single AF patients. The GWAS meta-analysis results indicate that recurrent AF patients seem to have a greater genetic risk of recurrent events and the PheWAS-analysis and gene co-expression network analyses highlight differences in the diseases and genetic functions associated to the set of SNPs and genes for to each group. In contrast, we find that for MI patients experiencing single events are significantly younger and have a better baseline health, yet they exhibit higher genetic risk. The GWAS meta-analysis identifies mostly genetic regions uniquely associated with single MI, and the PheWAS-analysis and gene co-expression networks support the genetic differences between these two groups.

## 1 INTRODUCTION

Myocardial infarction (MI) and atrial fibrillation (AF) are two major prevalent cardiovascular diseases. AF is an established risk factor for several other cardiovascular diseases, where the severity and mortality risk increase rapidly with relapse of AF. To treat and prevent new events of AF, medical and interventional therapy are available as treatments in order to normalize rythm or to stabilize the heart rate. AF ablation has become the leading clinical treatment, but with varying success rate, where 20% to 40% of the patients require new treatment (1). Similarly, MI is a severe heart diagnosis associated with high mortality rates. Upon survival, the heart is most likely weakened, making the patient vulnerable for other diseases. In fact, 33% of the patients experiencing MI die within a year (all death causes) (2). On the other hand, some patients experience only one event of MI and have a normal and healthy life afterwards. Most MI patients undergo cardiac catheterization and percutaneous coronary interventions in the acute phase, while medical therapy targeting clotting of blood and lipids as well as life-style interventions, are provided to reduce recurrent events. Still, a significant proportion of individuals suffers from relapse of MI.

Many genetic studies have been conducted to identify genetic variants that likely affect the risk of AF (3, 4) and MI (5, 6). While many variants have been identified and replicated in other studies, these variants were identified when comparing all included cases of AF or MI with healthy controls. Some studies have been conducted with the aim of understanding the genetic of patients experiencing recurrent events of AF (1, 7, 8, 9, 10) and MI (11, 12, 13). These are, however, mostly focused on either (A) response after treatment, or (B) the genetic effects on recurrence from known AF/MI variants or genes. Little effort has been made regarding the comparison of genetics between patients experiencing a single event versus patients experiencing recurrent events.

To our knowledge, no studies have been conducted with a full GWAS-analysis comparing recurrent to single events of AF and MI. In the presented study, we investigate if there are statistically significant genetic differences between patients experiencing recurrent events (defined as two or more events) of AF or MI compared to patients only experiencing a single event. Note that, we perform a general comparison without separating cases based on type of possible treatment after the first AF or MI event. By this, we perform a general comparison between single and recurrent events of AF and MI, to identify if there are any genetic differences between patients with single events to patients with recurrent events.

## 2 METHODS

### 2.1 Cohorts

#### 2.1.1 The HUNT study

The Trøndelag Health study (HUNT) is a health related population based longitudinal study, based on four rounds of data collection: HUNT1 (1984–1986), HUNT2 (1995–1997), HUNT3 (2006–2008) and HUNT4 (2017–2019). With a unique database covering clinical measurements, questionnaire data and biological samples from roughly 230, 000 of Trøndelag counties’ inhabitants from 1984 onward, it is one of the largest health studied ever performed (14). A great benefit of the HUNT study is the link with other health related registries through the Norwegian unique personal identification number. Such health registries include hospital and general practitioner registries, cancer registries, cause of death registries, and the prescription database.

In the current study, genotype data for 69, 621 participants from HUNT2 and HUNT3 were used, and these were linked to questionnaire data and clinical measurements from HUNT1, HUNT2, and HUNT3, regional hospital records, Nord-Trøndelag Hospital Trust (HNT), and the Norwegian Cause of Death registry (COD). The HNT register contains all ICD9 and ICD10-codes for registered hospital visits of these participants in the time period of August 1987 to April 2017. The COD registry spans the same time period, with registered ICD9 and ICD10-codes for the primary and secondary causes of death.

#### 2.1.2 The United Kingdom Biobank

The United Kingdom Biobank (UKBB) is a health related population based study consisting of roughly 500, 000 middle aged UK inhabitants from across the country. Sampling of the participants took place during 2006-2010, where questionnaires, clinical measurements, and biological samples were collected. Similar to the HUNT study, it is also linked to electronic health records, containing information about the participants hospital, general practitioner, and death records with ICD9 and ICD10 diagnose codes (15). Genotyped data are available for more than 480, 000 participants, and in the current study, we use these data together with relevant questionnaire data, clinical measurements, and hospital and death records for the genotyped participants (of European ancestry). The hospital records span the time period from December 1992 to September 2021. The death registry spans the same period, starting from 2006.

### 2.2 Genotyping and imputation

Genotyping and imputation for the HUNT and UKBB participants have been described elsewhere (16, 17). In short, genotyping was performed using one of three Illumina HumanCoreExome arrays: 12 v.1.0, 12 v.1.1 with custom content (UM HUNT Biobank v1.0) according to standard protocols for the HUNT participants, and standard protocols for Affymetrix Applied Biosystems UK BiLEVE Axiom or Applied Biosystems UK Biobank Axiom array for the UKBB participants. Standard quality control was performed for the HUNT genotyping, as well as a UKBB-specific quality control for the UKBB genotyping. Imputation in HUNT was performed using 2, 202 whole-genome sequenced samples from HUNT together with the Haplotype Reference Consortium (HRC) reference panel (18, 19), resulting in 25 million genetic markers. For UKBB, the HRC and UK10K+1000 Genomes reference panel was used, resulting in 90 million variants.

### 2.3 Definitions of traits and outcomes

Hospital records of HUNT and UKBB participants were used to determine cases of MI and AF, and the number of events for each participant. An event of MI is defined as having a registered diagnosis of ICD10:I21-I24 or ICD9:410. An event of AF is similarly defined with registered diagnosis of ICD10:I48 or ICD9:427.3.

In both records, each diagnosis is registered as a main or bi-diagnosis (denoted as first and second diagnosis in UKBB), and we consider them both when determining the number of events for each participant. Considering only the main diagnosis and filtering out bi-diagnosis can cause errors in addition to significantly reducing the amount of data. A bi-diagnosis can be interpreted in several ways: One could have the case where a person is admitted to the hospital with a main diagnosis, and additional diagnoses are detected and registered at this first visit. These will be registered as bi-diagnoses, even though they are new diagnoses and could have been recorded as main diagnoses if they were the only disease and the reason for the hospital visit that day. A different example could be that the doctor could inferred from previous records that other diseases (such as AF and MI) are relevant diagnoses for the main diagnosis and record these as bi-diagnoses, even though these are not new events. Due to the different ways of reporting bi-diagnosis, we apply some selective filtering in order to separate single events from recurring events of MI and AF.

A first event is defined as the first visit where this diagnosis is listed in the records, either as a main or a bi-diagnose. A second event is defined if there is more than one month time delay between the initial and the following events while one of the following two criteria are fulfilled: (i) the current event is a main diagnosis, (ii) the current event is a bi-diagnosis and the only diagnosis given that day (indicating that this is indeed a new event reported on this day). Following events are similarly registered as the second event, with a minimum one month time delay to the previously defined event. Except for the case of selecting the main diagnoses events exclusively, this is the only event definition that ensures the selection of new events.

As we are interested in contrasting patients with recurring events with relapse-free patients, we need to establish that the patients with only a single recorded event is not identified as such by the fact that they died before they experienced a new event. Identifying in the HUNT data that 80% of secondary events happened within a five year span for AF and seven year span for MI, we filter out single event participants who have died either due to the phenotype or within five or seven years after the AF or MI event, respectively (corresponding values were found in UKBB). Also, we take notice of the censoring dates of *06-04-2017* and *12-11-2021* for HUNT and UKBB respectively, and filter out participants registered with only one event of AF or MI, where their last AF or MI event is less than five and seven years prior to these censoring dates.

The four trait groups are from here denoted as single AF/MI: participants that experience only one event of AF/MI, and recurrent AF/MI: participants that experience more than one event of AF/MI, while satisfying the conditions specified above.

Baseline and clinical characteristics, as well as information about other relevant diseases identified with the participants, were taken from HUNT and UKBB hospital records, questionnaires, and clinical measurements. Participants were defined to have diabetes and/or hypertension if they have ever been registered with the ICD-codes ICD10:E10-E14 or ICD9:250 for diabetes and/or ICD10:I10-I15 (excluding I11.0), ICD9:401-405 for hypertension. The smoking variable is based on the HUNT question: “Have you ever smoked? (Yes/No)”, where the latest possible HUNT participation were used for each patient. The corresponding variable in UKBB reads “Ever smoked (Yes/No)” which was constructed upon sampling. BMI, cholesterol, systolic and diastolic blood pressure were measured in HUNT2 and HUNT3, where the HUNT study closest prior to the disease event were used. Corresponding variables were measured upon sampling in the UKBB. Student’s t-test for continuous variables and Fishers’ exact test for binary variables were used to test differences of the characteristics (age, diabetes, hypertension, BMI, smoking, cholesterol, systolic, and diastolic blood pressure) between groups of single versus recurrent events. Test statistics with Bonferroni-adjusted *p*-values (*p <* 0.05*/*8 = 6.25 *·* 10*^−^*^3^) were considered as significant findings.

### 2.4 GWAS meta-analysis

To identify genetic factors associated with single or recurrent events of AF and MI, we conducted three GWAS analyses for each trait in both cohorts separately: (i) patients with single events versus healthy controls, (ii) patients with recurrent events versus healthy controls, and (iii) patients with recurrent events versus patients with single events. As a control, we also conducted a GWAS analysis in each cohort with all cases of each disease against healthy controls. Healthy controls were defined as participants with no registered events of AF and MI, respectively. Variants with minor allele count (MAC)*<* 3 and an imputation score *<* 0.3 were excluded from all GWAS results. Participants with non-European recent ancestry were excluded from the analyses in UKBB (all genotyped HUNT participants are of European ancestry). Association analyses were performed with SAIGE, using a generalized linear mixed model adjusting for relatedness and unbalanced case-control ratios (20). Birth year, gender, batch/chip, and the first four principal components were added as covariates in the models. Genomic variants with minor allele frequency (MAF) *>* 1% in one or both studies were included in the meta-analysis.

From the eight GWAS analyses (three for AF and three for MI, and one control for each disease) performed for both the HUNT population and the UKBB population, we performed eight fixed-effect inverse variance weighted (IVW) meta-analyses using METAL (21). In METAL, each variant is assigned a new effect size as a sum of each study’s effect size weighted by the corresponding study variance. The *p*-values in the meta-analysis are calculated based on the *Z* statistic given by the new effect sizes and standard errors. Variants reaching genome-wide significance (*p*-values *<* 5 *·* 10*^−^*^8^) from the *Z* statistic were considered as significant findings. Annotations of the significant SNPs, identification of nearest genes, and search for nearby SNPs associated with relevant traits were performed with the FUMA platform and the GWAS catalog (22, 23). Variants were considered to be in the same genetic region if they were less than 500 kb apart, and genetic regions denoted as shared for both the single and recurrent events meta-analysis were either consisting of the same SNPs or SNPs within the same genetic region.

### 2.5 Phenome Wide Association Studies (PheWAS)

Phenome Wide Association Studies (PheWAS) were performed on all SNPs from the meta-analyses reaching genome-wide significance. From the comprehensive Pan UKBB resource (24), we collected results from GWAS conducted on 1, 326 phenocodes, and we identified the effect of each of our SNPs of interest on each phenocode. All GWAS results from the Pan UKBB are based on UKBB participants, and we selected results for European ancestry exclusively. For each set of SNPs (identified in common or specifically for either single or recurrent AF/MI), phenotypes with a *p*-value less than 0.05*/*(1326 *× n_set_*), where *n_set_* is the number of SNPs in the set, were considered as significant associations. For simplicity, only the SNP with the lowest *p*-value for each phenotype were selected among each set of SNPs.

### 2.6 Gene function and network analyses

The sets of nearest genes to the SNPs identified through the GWAS analyses as common or unique to either recurrent or single AF/MI events (in total six sets) were analyzed for tissue specificity (differentially expressed gene sets in each tissue). We employed both FUMA(22) and gene ontology enrichment using the Fishers Exact over-representation test in PANTHER (protein annotation through evolutionary relationship) (25). Here, biological processes with false discovery rate (FDR) adjusted for multiple testing *<* 0.05 were considered as functionally enriched for the gene set. To further investigate processes connected to these genes, we performed gene co-expression network analysis (26, 27, 28), where the hypothesis is that highly correlated genes have a regulatory relationship or similar response in a condition (29). Using the identified gene sets as target genes in an egocentric gene co-expression network analysis, we generate a network from the shared neighborhoods among the closest neighbour-genes of each target gene (in the gene) set and inspect the gene functions in the network.

Creating these egocentric networks involves several steps. First, using gene expression data from GTEx v.8(30) (https://www.gtexportal.org) gene co-expression networks for seven tissue sub-types from heart, muscle skeletal, artery, and kidney (GTEx Analysis v8 eQTL expression matrices.tar: *Heart Atrial Appendage, Heart Left Ventricle, Muscle Skeletal, Artery Aorta, Artery Coronary, Artery Tibial* and *Kidney Cortex*) were created. Since co-expression patterns may vary in different tissues (29), a separate network was created for each tissue. Following the WGCNA-approach (31), the link weight (strength of co-expression) between each pair of genes (*i* and *j*) were defined by the weighted topological overlap (*wTO*):

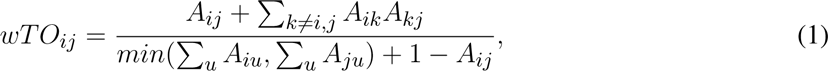

where *A_ij_* = *|cor*(*i, j*)*|*^6^ is the absolute Pearson correlation of the gene expressions raised to a power 6 to emphasize the strongest correlations. The resulting gene co-expression network is then an all-to-all network where pairs of genes with high *wTO*-link weights represent strong connections between the genes and their topological neighbourhood. Only the 15% strongest links from each tissue were included in the following analysis (still leaving about 30 million links), to avoid inclusion of genes based on weak (and likely spurious) connections.

Next, for each of the seven tissues, egocentric networks for each target gene were extracted from the co-expression networks. The egocentric networks were filtered to include only the 25 genes with the strongest *wTO*-link weights with each target gene. By merging and further reducing the complexity of the networks, the 25 strongest linked genes to each target gene across all tissues were selected in the final network. Here, we weighted the link strengths using 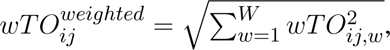, where *W* is the number of tissues in which the linked gene is among the 25 strongest linked gene to the target gene, and *wTO_ij,w_*, is the corresponding *wTO*-link weight in tissue *w*.

The final six sets of egocentric networks for target genes identified as common or as unique for the single or recurrent AF/MI events, were analyzed with the *igraph* R-package (32, 33). Shared neighbouring genes were defined as genes linked to two or more of the target genes. The set of shared neighbourhood genes for each network was plotted separately with Cytoscape (v. 3.8.1) (34) and gene ontology enrichment of these gene sets were obtained through the PANTHER over-representation test (25).

## 3 RESULTS

### 3.1 Characteristics of trait groups

Among the genotyped participants with European ancestry included in this study, there are 7, 127 and 29, 330 hospital patients registered with AF in HUNT and UKBB, respectively. With the filtering described in the methods section, we identify 1, 425 HUNT and 9, 561 UKBB patients with single AF events, and 2, 267 HUNT and 7, 267 UKBB patients with recurrent AF events. Correspondingly, 5, 805 HUNT and 14, 592 UKBB participants are registered with MI events, and 1, 651 HUNT and 6, 584 UKBB patients are identified with single MI events, and 1, 615 HUNT and 1, 615 UKBB patients are identified with recurrent MI events.

Baseline and clinical characteristics of these patients are presented in Tab. 1. Comparing the two AF groups in HUNT, there seems to be a tendency that the group experiencing single AF events are older with worse health conditions, having a higher percentage of diabetes and hypertension and being significantly older (adjusted *p*-value 2 *·* 10*^−^*^10^) with significantly higher levels of cholesterol and systolic blood pressure (adjusted *p*-values 6.1 *·* 10*^−^*^6^ and 5 *·* 10*^−^*^4^). Comparing the AF groups in UKBB, we see the same tendency with significantly higher age (adjusted *p*-value *<* 10*^−^*^16^) and somewhat higher levels of BMI and systolic blood pressure for the single AF event group compared to the recurrent AF group. Comparing the MI groups in HUNT, we see the reverse trends, where patients experiencing recurrent MI events are significantly older (adjusted *p*-value *<* 10*^−^*^16^) with significantly higher percentages of diabetes and hypertension (adjusted *p*-values 8.8 *·* 10*^−^*^5^ and 7.6 *·* 10*^−^*^3^) and significantly higher levels of cholesterol and systolic blood pressure (adjusted *p*-values 5.4 *·* 10*^−^*^6^ and 1.4 *·* 10*^−^*^5^). There is also a tendency of higher BMI and diastolic blood pressure for this group. The same trends are found in the UKBB MI groups, with significantly higher age, BMI and percentage of diabetes and hypertension (adjusted *p*-values 6 *·* 10*^−^*^14^, 3.9 *·* 10*^−^*^5^, *<* 10*^−^*^16^ and *<* 10*^−^*^16^) for the recurrent MI events group compared to the single MI events group. There is also a tendency towards higher levels of systolic blood pressure for the UKBB single MI events group.

**Table 1.**
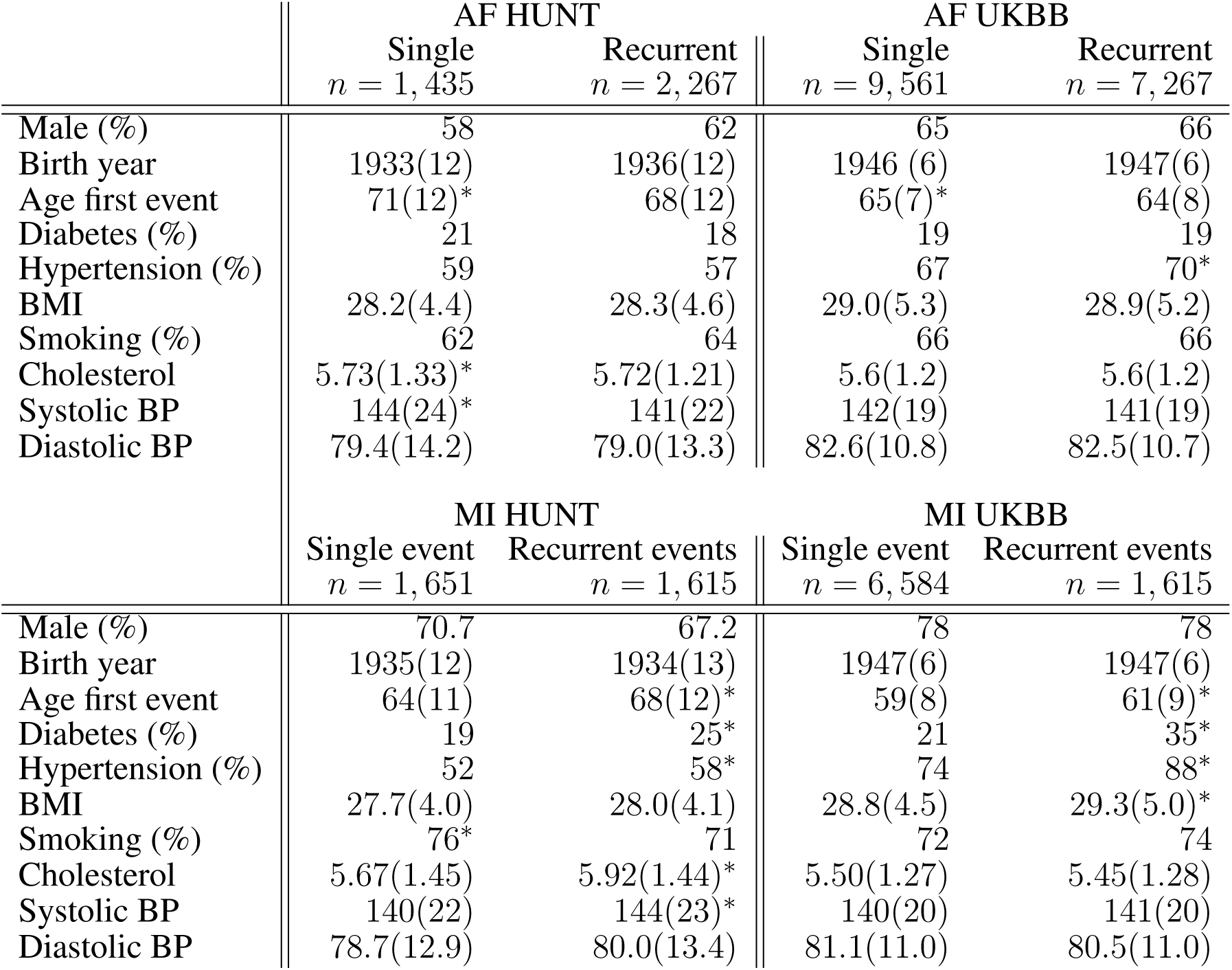
Characteristics of sample groups of single and recurrent events of AF and MI in the HUNT and UKBB population. Continuous variables are presented as mean and (sd), and dichotomous variables are reported in percentages. Starred values indicate a significant difference between the single and recurrent group, where the starred value is marking the larger value.

|  | AF HUNT |  | AF UKBB |  |
| --- | --- | --- | --- | --- |
|  | Single<br>*n* = 1,435 | Recurrent<br>*n* = 2,267 | Single<br>*n* = 9,561 | Recurrent<br>*n* = 7,267 |
| Male (%) | 58 | 62 | 65 | 66 |
| Birth year | 1933(12) | 1936(12) | 1946(6) | 1947(6) |
| Age first event | 71(12)* | 68(12) | 65(7)* | 64(8) |
| Diabetes (%) | 21 | 18 | 19 | 19 |
| Hypertension (%) | 59 | 57 | 67 | 70* |
| BMI | 28.2(4.4) | 28.3(4.6) | 29.0(5.3) | 28.9(5.2) |
| Smoking (%) | 62 | 64 | 66 | 66 |
| Cholesterol | 5.73(1.33)* | 5.72(1.21) | 5.6(1.2) | 5.6(1.2) |
| Systolic BP | 144(24)* | 141(22) | 142(19) | 141(19) |
| Diastolic BP | 79.4(14.2) | 79.0(13.3) | 82.6(10.8) | 82.5(10.7) |
|  | MI HUNT |  | MI UKBB |  |
|  | Single event<br>*n* = 1,651 | Recurrent events<br>*n* = 1,615 | Single event<br>*n* = 6,584 | Recurrent events<br>*n* = 1,615 |
| Male (%) | 70.7 | 67.2 | 78 | 78 |
| Birth year | 1935(12) | 1934(13) | 1947(6) | 1947(6) |
| Age first event | 64(11) | 68(12)* | 59(8) | 61(9)* |
| Diabetes (%) | 19 | 25* | 21 | 35* |
| Hypertension (%) | 52 | 58* | 74 | 88* |
| BMI | 27.7(4.0) | 28.0(4.1) | 28.8(4.5) | 29.3(5.0)* |
| Smoking (%) | 76* | 71 | 72 | 74 |
| Cholesterol | 5.67(1.45) | 5.92(1.44)* | 5.50(1.27) | 5.45(1.28) |
| Systolic BP | 140(22) | 144(23)* | 140(20) | 141(20) |
| Diastolic BP | 78.7(12.9) | 80.0(13.4) | 81.1(11.0) | 80.5(11.0) |

In total, these are indications of patients experiencing only single AF events being older at their first event and with worse health condition and lifestyle factors compared to the recurrent AF cases. We therefore hypothesise that single AF events could be more likely driven by age and other lifestyle factors, while there are genetic factors driving recurrent AF events. For MI, the characteristics point in the opposite direction, where patients experiencing recurrent MI events are older and generally of worse health conditions and lifestyle factors compared to patients with only one MI event and surviving it. For MI, we therefore formulate two hypothesis: (i) that recurrent MI events are driven by the age at first event and worse health conditions, while single MI events are driven by genetic factors, or (ii) that both single and recurrent MI are driven by the same genetic factors, but recurring MI are due to higher age and other lifestyle factors affecting the risk of obtaining new MI events.

### 3.2 Genetic differences

In the following, we explore our hypothesis (defined above) for AF and MI by investigating genetic differences between the groups identified through the GWAS meta-analyses.

#### 3.2.1 Genetic differences in AF

To test our hypothesis that patients experiencing recurrent AF events are more genetically susceptible than patients experiencing single AF, we perform three GWAS-meta analyses (see Methods). The GWAS meta-analysis comparing single to recurrent AF events found no regions with significantly different effects. Some SNPs were identified genome-wide significant in the HUNT population, but these were rare variants (MAF *≤* 0.2%) and removed through filtering prior to the META-analysis. Comparing the GWAS meta-analyses of each group against healthy controls (Tab. 2 and Fig. 1), we find that 18 regions are specifically associated with recurrent AF events, 2 are specifically associated with single AF events and 16 are identified in both GWAS investigations. Many regions comprise multiple SNPs that exhibit significant effects in only one of the study groups. Five regions identified in the recurrent AF GWAS study consist of only one SNP, yet these are identified with similarly strong effects in both the HUNT and the UKBB studies, indicating a genuine association. Regional plots of the single SNP hits uniquely associated with recurrent AF are shown in Additional File 1, Fig.S15-S19. The presented results show that over half of the identified regions are specifically linked to single or recurrent AF, supporting the hypothesis that patients who have experienced recurrent AF events are genetically more susceptible than those who have only experienced one event and survived it.

**Figure 1.**
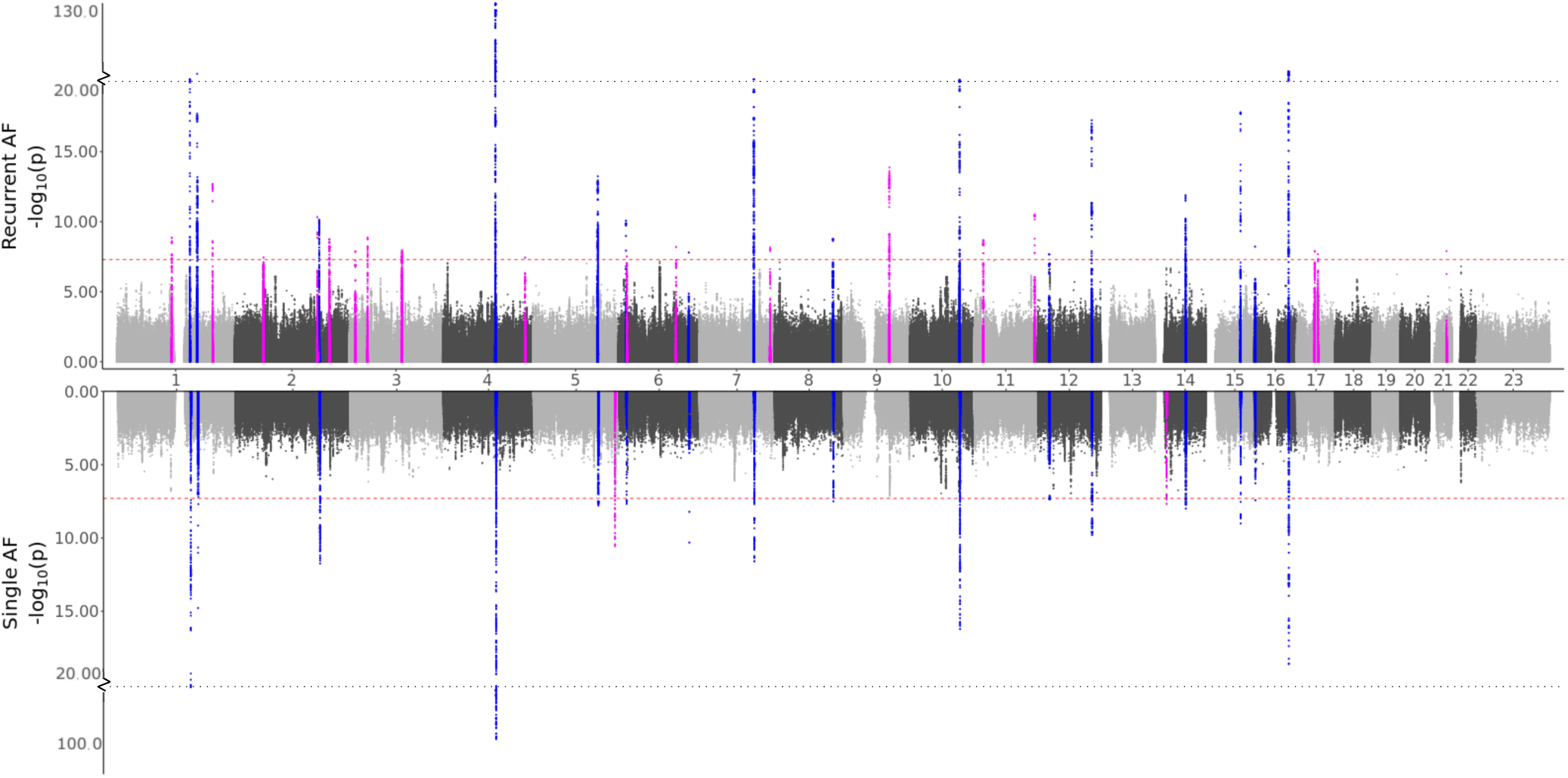
GWAS meta analysis results for Atrial Fibrillation (AF). Top: Comparing recurrent AF patients to AF-free controls. Bottom: Comparing single AF patients to AF-free controls. Blue colored spikes represent regions of SNPs found to be statistically significant in both GWAS studies (common), while magenta spikes represent statistically significant regions of SNPs specifically associated with the given AF group.

**Table 2.** Atrial Fibrillation (AF) variants found to be significant in the GWAS meta analysis. The lead variant (lowest *p*-value) for each independent region is listed. The “AFfull” column describes if the variant (or any of the significant variants in this region) is also identified in the full AF GWAS meta analysis. The “AF Study” column shows in which of the studies the variant/region was found to be significant (”common” meaning significant in both the single and the recurrent GWAS meta analysis). The “Known” column reports if this region is previously known for AF. The subsequent columns are “rsID,” position, nearest gene, and the function of the lead SNP as well as effect size, standard error, and *p*-value from the meta-analysis. “Dir” corresponds to the direction of the effect in the HUNT and UKBB GWAS meta-analysis respectively (an entry ofmeaning not included in the meta-analysis - see Additional File 4 for allele frequencies), and “Nsnps” shows the number of significant SNPs in the region.

| AFull | AF Study | Known | rsID | Chr:pos_Ref/Alt | Gene | Function | Effect | StdErr | Pvalue | Dir | Nsnps |
| --- | --- | --- | --- | --- | --- | --- | --- | --- | --- | --- | --- |
| Yes | Common | Yes | rs6691463 | 1:154814538_C/G | KCNN3 | intronic | -0.16 | 0.015 | $3.5 \times 10^{-24}$ | -- | 166 |
| Yes | Common | Yes | rs72700114 | 1:170193825_G/C | NTMT2 | intergenic | 0.36 | 0.031 | $9.1 \times 10^{-32}$ | ++ | 239 |
| Yes | Common | Yes | rs1429094 | 2:179515774_A/G | TTN | intergenic | -0.14 | 0.02 | $1.8 \times 10^{-12}$ | -- | 64 |
| Yes | Common | Yes | rs6843082 | 4:111718067_G/A | PITX2 | intergenic | -0.5 | 0.021 | $2.4 \times 10^{-132}$ | -- | 553 |
| Yes | Common | Yes | rs678897 | 5:137441065_G/C | NME5 | intergenic | 0.13 | 0.017 | $5.7 \times 10^{-14}$ | ++ | 229 |
| Yes | Common | Yes | rs112899072 | 6:16417852_C/G | ATXN1 | intronic | 0.15 | 0.023 | $8.6 \times 10^{-11}$ | ++ | 10 |
| Yes | Common | Yes | rs117984853 | 6:149399100_G/T | UST | downstream | 0.16 | 0.025 | $4.9 \times 10^{-11}$ | ++ | 2 |
| Yes | Common | Yes | rs11773845 | 7:116191301_C/A | CAV1 | intergenic | 0.16 | 0.016 | $4 \times 10^{-24}$ | ++ | 218 |
| Yes | Common | Yes | rs17337621 | 8:124542519_G/C | FBXO32 | intronic | 0.19 | 0.032 | $1.7 \times 10^{-9}$ | ++ | 5 |
| Yes | Common | Yes | rs1389189 | 10:105486077_A/G | SH3PXD2A | intronic | -0.14 | 0.017 | $6.1 \times 10^{-17}$ | -- | 122 |
| Yes | Common | Yes | rs137913153 | 12:24776752_A/G | SOX5 | intergenic | 0.11 | 0.02 | $4.2 \times 10^{-8}$ | ++ | 8 |
| Yes | Common | Yes | rs883079 | 12:114793240_C/T | TBX5 | UTR3 | 0.15 | 0.017 | $5.8 \times 10^{-18}$ | ++ | 67 |
| Yes | Common | Yes | rs1152591 | 14:64680848_A/G | SYNE2 | intronic | 0.11 | 0.015 | $1.3 \times 10^{-12}$ | ++ | 67 |
| Yes | Common | Yes | rs7172038 | 15:73667255_T/G | HCN4 | intergenic | -0.18 | 0.021 | $1.6 \times 10^{-18}$ | -- | 44 |
| Yes | Common | Yes | rs140185678 | 16:2003016_G/A | RPL3L | exonic | 0.25 | 0.043 | $6.1 \times 10^{-9}$ | ++ | 1 |
| Yes | Common | Yes | rs4499262 | 16:73059159_C/A | ZFHX3 | intronic | 0.26 | 0.021 | $2.5 \times 10^{-35}$ | ++ | 108 |
| Yes | Recurrent | Yes | rs9428227 | 1:116309200_C/T | CASQ2 | intronic | 0.093 | 0.015 | $1.3 \times 10^{-9}$ | ++ | 24 |
| Yes | Recurrent | Yes | rs2270543 | 1:203030685_T/C | PPFIA4 | intronic | 0.11 | 0.015 | $2 \times 10^{-13}$ | ++ | 26 |
| Yes | Recurrent | Yes | rs12463885 | 2:61457996_A/C | USP34 | intronic | 0.086 | 0.016 | $3.6 \times 10^{-8}$ | ++ | 1 |
| Yes | Recurrent | Yes | rs56181519 | 2:175555714_C/T | WIPF1 | intergenic | -0.12 | 0.018 | $4.8 \times 10^{-11}$ | -- | 4 |
| Yes | Recurrent | Yes | rs7605146 | 2:201183888_G/A | SPATS2L | intronic | 0.094 | 0.016 | $1.8 \times 10^{-9}$ | ++ | 40 |
| Yes | Recurrent | Yes | rs4642101 | 3:12842223_T/G | CAND2 | intronic | -0.091 | 0.016 | $1.3 \times 10^{-8}$ | -- | 2 |
| Yes | Recurrent | Yes | rs6790396 | 3:38771925_C/G | SCN10A | intronic | -0.095 | 0.016 | $1.4 \times 10^{-9}$ | -- | 16 |
| Yes | Recurrent | Yes | rs73228569 | 3:111614052_T/C | PHLDB2 | intronic | 0.12 | 0.021 | $1.5 \times 10^{-8}$ | ++ | 34 |
| Yes | Recurrent | Yes | rs12646050 | 4:174634261_A/G | RANP6 | intergenic | 0.14 | 0.026 | $3.7 \times 10^{-8}$ | ++ | 1 |
| Yes | Recurrent | Yes | rs34969716 | 6:18210109_G/A | KDM1B | intronic | 0.095 | 0.017 | $3.1 \times 10^{-8}$ | ++ | 1 |
| Yes | Recurrent | Yes | rs2684249 | 6:122392511_T/C | HSF2 | intergenic | 0.09 | 0.016 | $6.4 \times 10^{-9}$ | ++ | 1 |
| No | Recurrent | Yes | rs758890 | 7:150655643_G/A | KCNH2 | intronic | -0.094 | 0.016 | $7 \times 10^{-9}$ | -- | 5 |
| Yes | Recurrent | Yes | rs10993463 | 9:97807233_C/T | AOPEP | intronic | 0.12 | 0.016 | $1.3 \times 10^{-14}$ | ++ | 88 |
| Yes | Recurrent | Yes | rs2568119 | 11:20004957_G/A | NAV2 | intronic | 0.11 | 0.018 | $2 \times 10^{-9}$ | ++ | 26 |
| Yes | Recurrent | Yes | rs3765618 | 11:128769876_C/G | KCNJ5 | UTR3 | -0.18 | 0.027 | $3.2 \times 10^{-11}$ | -- | 12 |
| Yes | Recurrent | Yes | rs12944882 | 17:37983492_T/C | IKZF3 | intronic | 0.088 | 0.015 | $1.2 \times 10^{-8}$ | ++ | 2 |
| Yes | Recurrent | Yes | rs17608766 | 17:45013271_T/C | GOSR2 | UTR3 | -0.12 | 0.022 | $2.1 \times 10^{-8}$ | -- | 2 |
| Yes | Recurrent | Yes | rs2834618 | 21:36119111_T/G | CLIC6 | intergenic | 0.15 | 0.026 | $1.2 \times 10^{-8}$ | ++ | 1 |
| Yes | Single | Yes | rs1223535129 | 5:172664353_CG/C | NKX2-5 | intergenic | 0.11 | 0.017 | $2.7 \times 10^{-11}$ | ?+ | 31 |
| Yes | Single | Yes | rs28631169 | 14:23888183_C/T | MYH7 | intronic | 0.1 | 0.018 | $2.1 \times 10^{-8}$ | ++ | 2 |

All regions had previously been associated with AF, and all regions except one (the region at chromosome 7 in the KCNH2 gene) were identified in the full AF GWAS meta-analysis by comparing all AF cases against healthy controls. This indicates that all the regions identified as unique for single or recurrent AF (excluding the KCNH2 gene region) have an effect when compared to healthy controls, with the true effect being mainly or solely for patients experiencing single or recurrent AF. The five SNPs in the KCNH2 gene, however, are not detected in our full GWAS meta-analysis and therefore only show an effect for patients experiencing recurrent AF.

To our knowledge, only seven genes have been found to be associated with AF recurrence: SOX5, CAV1, EPHX2, ITGA9, SLC8A1, TBX5 and PITX2 (10, 8, 35, 1). Our findings show that regions proximate to the SOX5, CAV1, TBX5 and PITX2 genes are identified in both the single and the recurrent GWAS, yielding comparable effects. Thus, no differences in the impact of these regions between the two groups are evident. No regions were identified near the EPHX2, SCL8A1 and ITGA9 genes. Variants near the NAV2 and SCN10A genes have previously been tested for their effect in recurrent AF events without any significant findings (36, 35). In this study, we discovered 26 and 16 SNPs located within and nearby the NAV2 and SCN10A genes, respectively, that are exclusively associated with recurrent AF, suggesting that these SNPs have a distinct effect on recurrent AF patients compared to single AF cases.

Several of the genes listed in Tab.2 code for functions related to AF. Two of the genes listed as “Common” (KCNN3 and HCN4) and three genes identified uniquely for recurrent AF (SCN10A, KCNH2 and KCNJ5) are related to electrophysiological activity, coding for potassium and sodium channels. Other genes listed as “Common” in Tab. 2 code for functions directly linked to heart activity and AF (TTN, TBX5, SYNE2 and RPL3L) or indirectly linked through comorbidities (ATXN1, CAV1, SH3PXD2A and ZFHX3). Two of the recurrent AF genes also code for functions directly or indirectly linked to AF (CASQ2 and GOSR2), and some indicate a possible indirect link related to comorbidities, e.g. hypertension or malignancy (PPFIA4, USP34, WIPF1, SPATS2L, CAND2 and AOPEP). The two genes uniquely identified for single AF have been shown important for myocardial diseases and cardiac abnormalities, coding for functions found central in malformation in heart (NKX2-5) and myosin (MYH7).

#### 3.2.2 Genetic differences in MI

Based on the characteristics of the two MI groups, we formulate two hypotheses: (i) Single MI events may be driven by genetic factors and may be more susceptible to medical treatment, while recurrent MI events may be driven by higher age at the first event and other lifestyle factors, and (ii) Both single and recurrent MI are driven by the same genetic factors, but recurring MI events occur due to higher age and lifestyle factors. Testing for direct genetic differences between the two MI groups, the GWAS meta-analysis (comparing single to recurrent events) did not detect any regions with significant effects. When testing for genetic effects in each groups when compared to MI-free controls, the GWAS meta-analyses shown in Fig. 2 and Tab. 3 identified four regions that are in common for both groups, 24 regions that are specifically identified for the single event groups, and two regions that are unique for the recurrent events group. Hence, some genetic factors are common for both groups, but most identified genetic effects are unique to patients experiencing only one event of MI and surviving it. This results is in support of our first hypothesis.

**Figure 2.**
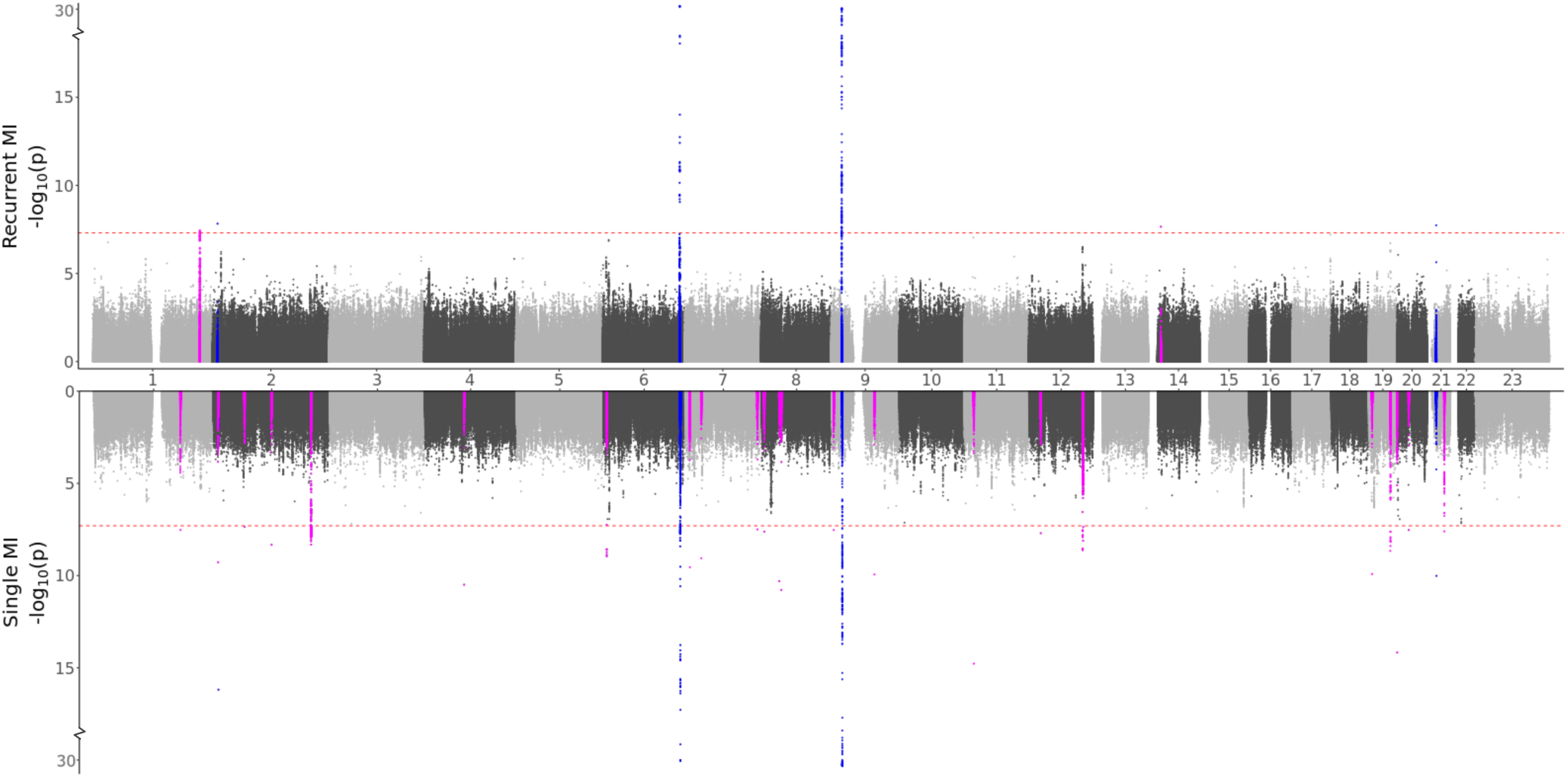
GWAS meta analysis results for Myocardial Infarction (MI). Top: Comparing recurrent MI patients to MI-free controls. Bottom: Comparing single MI patients to MI-free controls. Blue colored spikes represent regions of SNPs found to be statistically significant in both GWAS studies (common), while magenta spikes represents statistically significant regions of SNPs that are specifically associated with the given MI group.

**Table 3.**
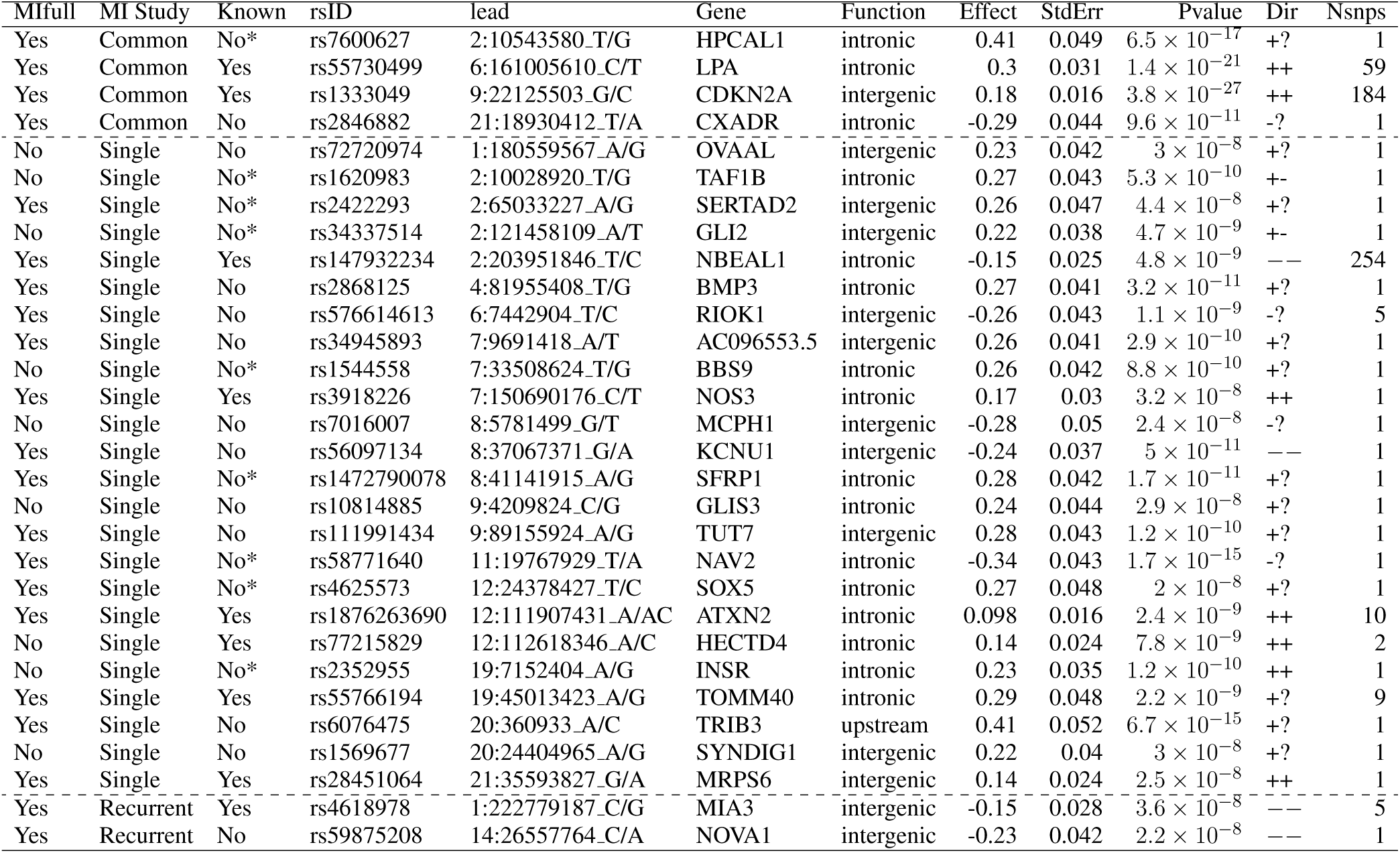
Myocardial Infarction (MI) variants that are significant in the GWAS meta analysis. The lead variant (lowest *p*-value) for each independent region is listed. The “MIfull” column describes if the given variant (or any of the significant variants in this region) is also identified in the full MI GWAS meta analysis. The “MI Study” column shows in which of our studies the variant/region was found to be significant (an entry of “common” means that it was present in both the single and the recurrent GWAS meta analysis). The “Known” column shows if this region is previously related to MI (”Yes”), a relevant CVD-trait (”No*”), or neither (”No”). The next columns are “rsID,” nearest gene, the function of the lead SNP as well as effect size, standard error, and *p*-value from the meta-analysis. “Dir” corresponds to the direction of the effect in the HUNT and UKBB GWAS meta-analysis respectively (a value “?” indicates that it was not included in the meta-analysis - see Additional File 4 for allele frequencies), and “Nsnps” shows the number of significant SNPs in the region.

Some distinct regions, including the SNPs in the NBEAL1 and ATXN2 genes for single MI events, and SNPs in the MIA3 gene for recurrent events, exhibit substantial effects for multiple SNPs in the region, with comparable effects in both the HUNT and UKBB populations. Several regions represent suggestive findings comprising only single SNPs and are only identified in the HUNT population (regional plots of the single SNP hits uniquely associated with single or recurrent MI are shown in Additional File 1, Fig.S20-S39). However, as shown in Additional File 4, these variants are not HUNT-specific since they are reported with relative high frequencies in the general European population. Hence, although they are rare in the UKBB population and thereby not included in the META-analysis, including a different European study population could validate or dispute the effect identified here. Also, many of these regions are well-known for MI, further indicating that these findings might be valid.

Comparing the GWAS meta-analysis of all MI cases to MI-free controls, we find that all four regions that were identified in common for single and recurrent MI (regions in or close to the genes HPCAL1, LPA, CDKN2A and CXADR) were also found in the full MI GWAS meta-analysis. The two regions that were specifically associated with recurrent MI events (regions close to the MIA3 and NOVA1 genes) were also identified in the full MI GWAS, but nine of the regions specifically associated with single MI were not detected in the full MI GWAS (regions in or close to the OVAAL, TAF1B, GLI2, BBS9, MCPH1, GLIS3, HECTD4, INSR and SYNDIG1). Hence, certain regions identified in the full MI GWAS are exclusively linked to either single or recurrent MI, and some regions are only observed when patients with single MI events are filtered out, emphasizing the need for dividing the MI groups.

Ten of the 24 regions specifically associated with single MI (the regions in or close to OVAAL, BMP3, RIOK1, AC096553.5, MCPH1, KCNU1, GLIS3, TUT7, TRIB3 and SYNDIG1) are novel findings for MI, and thus, have not previously been associated with CVD-related traits. While these regions consist of only a single SNP (with the exception of the five SNPs close to the RIOK1 gene) and are only found in the HUNT population (except for the SNP close to the KCNU1 gene), some of them are coding for similar functions to the identified previously known MI genes. Three of these genes are commonly related to malignancy (OVAAL, RIOK1, TUT7), similar to NBEAL1, in which we identify a known MI region consisting of 254 SNPs uniquely associated with single MI. Other genes are coding for proteins involved in calcium handling (BMP3, SYNDIG1) or associated with diabetes mellitus (GLIS3, TRIB3), which points toward a more aggressive development of atherosclerosis. Similarly, both ATXN2 and HECTD4, for which we identify known MI regions uniquely associated with single MI, are associated with diabetes mellitus. The two regions exclusively linked to recurrent MI events are both found with negative effects in both the HUNT and the UKBB population. The region close to the MIA3 gene has been associated with MI previously, while the single SNP close to the NOVA1 gene (also with possible relations to malignancy) is a novel finding. Taken together, these results highlight some interesting genes with a possible relation to MI. Further studies are needed to investigate if similar effects are found in other European and non-European populations and if these SNPs/genes link to MI and specifically link to either single or recurrent MI events.

### 3.3 Identification of additional phenotypes affected by SNPs through PheWas

To delve deeper into the genetic distinctions observed between single and recurrent AF and MI, we conducted a PheWAS (Phenome-Wide Association Study) analysis. This enabled us to pinpoint other phenotypes that are associated with the same set of SNPs that were designated as either common or unique for single and recurrent AF and MI.

Our PheWAS investigation of the SNPs identified as common for both single and recurrent AF revealed a total of 1, 903 SNPs which were linked with 236 phenocodes (shown in Fig. 3 and Additional File 2). Not surprisingly, the strongest associations were found for *Atrial fibrillation and flutter* and *Cardiac dysrhythmia* (*p*-value 10*^−^*^400^ and 10*^−^*^220^). Furthermore, we discerned robust associations to phenocodes related to *Appendiceal conditions* and *Coagulation defects*. Notably, the circulatory system category emerged as the predominant category, encompassing 54 phenocodes. This includes, but is not limited to, conditions such as *Phlebitis and thrombophlebitis, Sinoatrial node dysfunction (Bradycardia), Heart failure*, and *Hypertension*.

**Figure 3.**
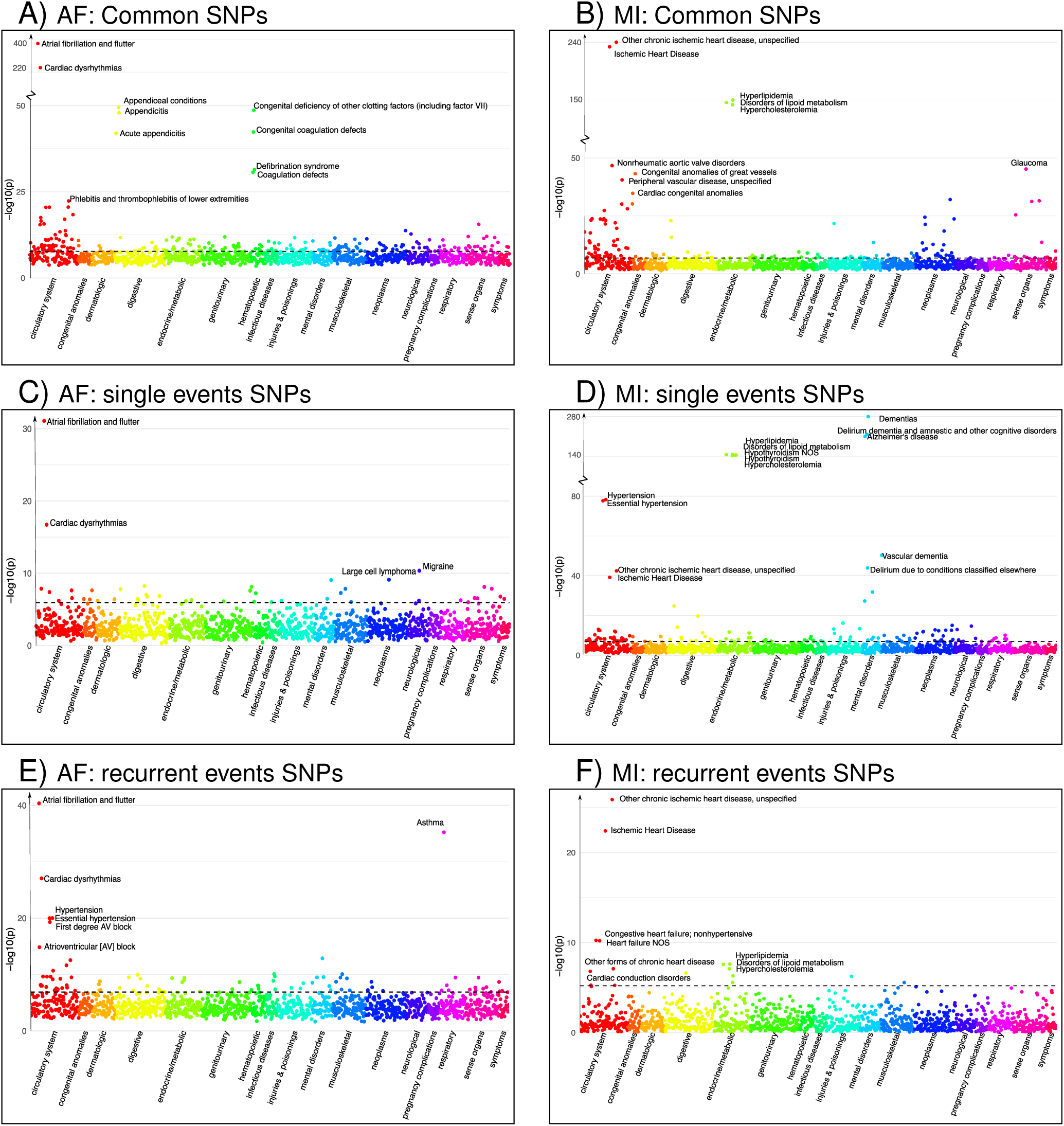
Phenocodes associated with each set of SNPs found for both single and recurrent AF/MI or uniquely for one of them. The *x*-axis shows each of the 1326 phenocodes sorted by phenocode category, and the *y*-axis shows the lowest *p*-value for the association between the phenocode and the SNPs in the set. The dotted line shows the threshold for significant associates, which vary according to the number of SNPs in each set. A) set of 1, 903 SNPs found in common for both single and recurrent AF. B) set of 245 SNPs found in common for both single and recurrent MI. C) set of 33 SNPs found uniquely for single AF events. D) set of 299 SNPs found uniquely for single MI events. E) set of 286 SNPs found uniquely for recurrent AF events. F) set of 6 SNPs found uniquely for recurrent MI events.

Intriguingly, the two identified regions specifically associated with single AF consist of 33 SNPs that exhibit significant association with 44 phenocodes (see Additional File 2). Not surprisingly, the strongest associations for these SNPs pertain to the phenocodes *Atrial fibrillation and flutter* and *Cardiac dysrhythmias*, the remaining 42 phenocodes span a diverse array of phenocode categories. These include not only *Migraine* and *Large cell lymphoma*, but also conditions such as *Arrhythmia (cardiac) NOS, Paroxysmal supraventricular tachycardia*, and *Cerebral atherosclerosis*.

In contrast, the 18 regions specifically associated with recurrent AF events consisting of 286 SNPs show significant association with 91 phenocodes (see Additional File 2), and a majority of the strong associations pertain to phenocodes of the circulatory system category. Again, the phenocode with the most potent associations are *Atrial fibrillation and flutter*, and *Cardiac dysrhythmias*. Additionally, these SNPs also display strong significant associations with *Asthma* and 27 phenocodes from the circulatory system, including conditions such as *Hypertension, Atrioventricular block, Cardiomyopathy, Heart failure, Ischemic heart disease, Cardiac arrest*, and *Palpitations*. Collectively, these results underscore genetic susceptibility disparities between patients experiencing single versus recurrent AF events. Specifically, SNPs specifically tied to recurrent AF are linked to a broad range of phenocodes related to the heart and circulatory system compared to SNPs specifically associated to single AF events.

Regarding MI, we identified four regions associated with both single and recurrent MI, comprising 245 SNPs that exhibit significant associations to 144 phenocodes (see Additional File 2). The most prominent associations are observed with *Ischemic heart disease* and *Hyperlipidemia* disorders. Additionally, numerous diseases within the circulatory system category, such as *Nonrheumatic aortic valve disorders, Peripheral vascular disease, Stricture of artery, Hypertension*, and *Heart valve disorders*, are also strongly associated.

The 24 regions specifically identified for single MI events consist of 299 SNPs that are associated with 128 phenocodes (see Additional File 2). These include *Ischemic heart disease, Hypertension*, and diseases of *Hyperlipidemia*. Additionally, there are strong associations with neurodegenerative disorders, such as *Dementia, Alzheimer’s*, and *Delirium*. These SNPs are furthermore linked with 33 phenocodes from the circulatory system category, highlighting conditions such as *Cerebral ischemia, Cardiac conduction disorders, Heart failure, Aortic valve disease*, and *Pulmonary heart disease*.

Notably, the two regions consisting of 6 SNPs specifically identified for recurrent MI were associated with a mere 16 phenocodes (see Additional File 2). While these included *Ischemic heart disease, Heart failure, Cardiac conduction disorders*, and diseases of *Hyperlipidemia*, they lacked the other 27 circulatory system disorders identified for the single MI SNPs. Once again, these findings emphazise the genetic differences between patients experiencing single and recurrent MI. SNPs specifically associated with single MI events appear to be associated with a broader and more diverse range of cardiovascular disorders compared to those solely linked to recurrent MI.

### 3.4 Gene sets and co-expression network neighbourhood

In our final analysis, we leverage multiple sets of gene expression data from the GTEx consortium (30) measured in tissue sub-types taken from heart, muscle, skeletal, artery, and kidney to generate gene co-expression networks (see Methods for details). Here, our expectation is that highly correlated genes have a regulatory relationship or similar response in a condition (29). Thus, this approach should uncover genes that display an expression profile that most closely links to the set of target genes found through our GWAS analyses, and we investigate their functions.

#### 3.4.1 AF associated genes in co-expression networks

Differential gene expression analysis of the 18 genes identified in recurrent AF (listed as Recurrent in Tab. 2) reveals a significant up-regulation of these genes in atrial appendage tissues from the heart. Furthermore, elevated expression levels are discerned in left ventricular heart, artery tibial, and muscle skeletal tissues (see Additional File 1, Fig. S9). Gene ontology analysis indicates that this set of genes is significantly enriched for cell-cell signaling involved in cardiac conduction, (Fold Enrichment (FE) *>* 100, FDR = 1.17 *×* 10*^−^*^2^), cardiac muscle cell action potential (FE = 70.03, FDR = 3.15 *×* 10*^−^*^2^), and regulation of heart rate (FE = 43.99, FDR = 1.62 *×* 10*^−^*^2^).

Following the co-expression analysis approach detailed in the Methods section, we find that 16 of the 18 recurrent AF genes show strong co-expression with other genes in heart, artery, kidney, and muscle skeletal tissues. Selecting the top-25 genes with the strongest connection to each of the 16 target genes, Fig. 4 A) shows that all of the 16 target genes are connected through 82 shared neighbouring genes (see Additional File 3), i.e. the 82 shared genes are among the top-25 strongest connections for two or more of the target genes. These 82 neighbouring genes are significantly enriched for a variety of biological processes, including acetyl-CoA biosynthetic process from pyruvate (FE*>* 100, FDR = 4.41 *×* 10*^−^*^3^), tricarboxylic acid cycle (FE= 54.70, FDR = 1.38 *×* 10*^−^*^6^), NLS-bearing protein import into nucleus (FE = 50.99, FDR = 1.93 *×* 10*^−^*^3^), inner mitochondrial membrane organization (FE = 32.73, FDR = 9.49 *×* 10*^−^*^4^), respiratory electron transport chain (FE = 10.72, FDR = 4.32 *×* 10*^−^*^2^), regulation of proteasomal protein catabolic process (FE = 7.49, FDR = 4.94 *×* 10*^−^*^2^), proteasome-mediated ubiquitin-dependent protein catabolic process (FE = 5.58, FDR= 3.79 *×* 10*^−^*^2^), and regulation of cellular catabolic process (FE = 4.01, FDR= 9.85 *×* 10*^−^*^3^).

**Figure 4.**
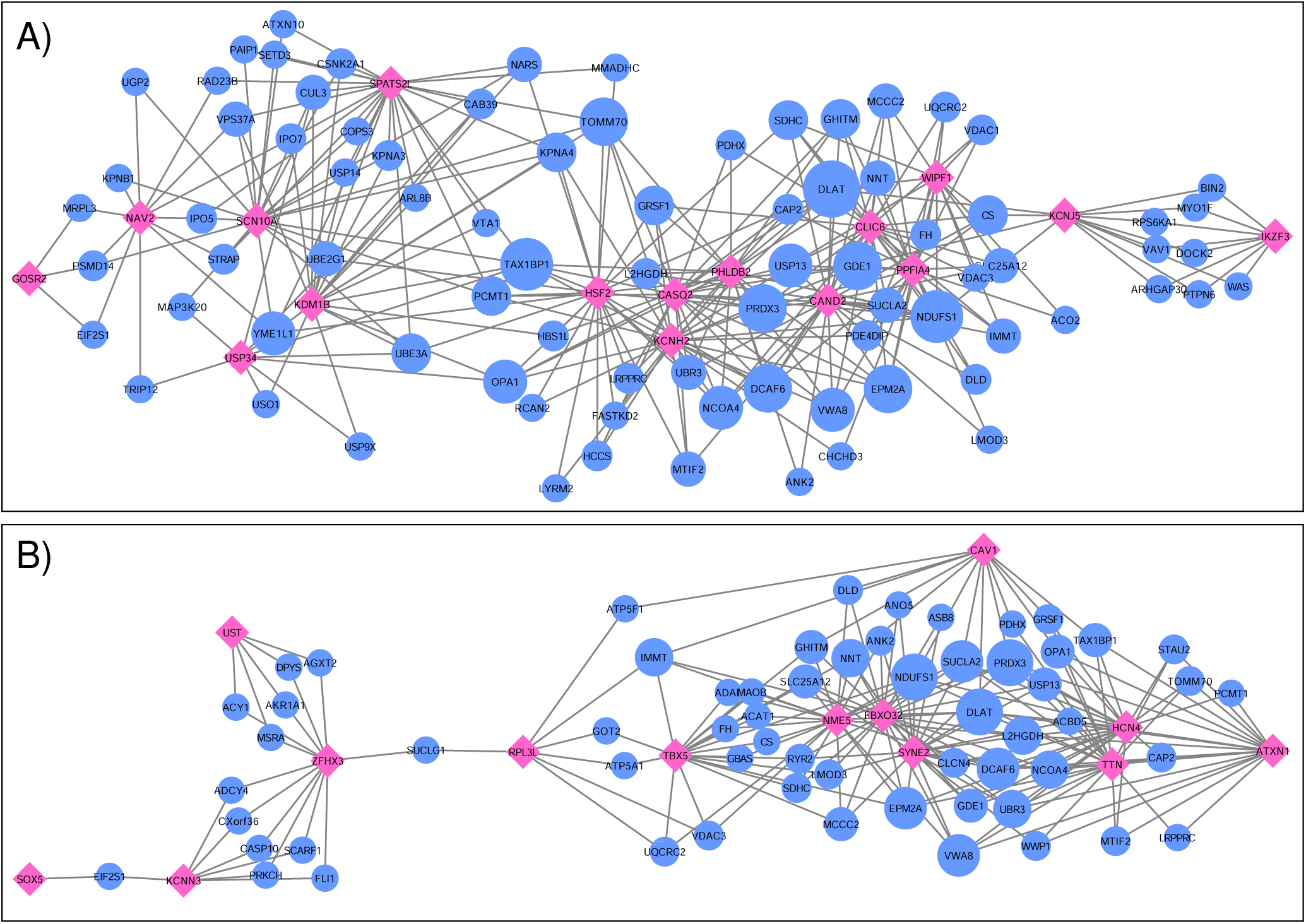
Networks showing the strongest shared neighbourhood of co-expressed genes for the GWAS (target) genes associated with A) recurrent AF uniquely and B) both single and recurrent AF. Pink diamond nodes represent the target genes and blue circular nodes represent the neighbouring genes. The sizes of the blue nodes are scaled according to their number of nearest neighbours in the network.

Focusing on the two genes specific to single AF events (listed as Single in Tab. 2), our analysis reveals that these genes shows significant up-regulation in left ventricle tissues of the heart and also high expression levels for atrial appendage tissues of the heart (see Additional File 1, Fig. S10). Gene ontology analysis confirms that these genes are closely linked to adult heart development (FE*>* 100, FDR = 7.77 *×* 10*^−^*^3^), ventricular cardiac muscle tissue morphogenesis (FE*>* 100, FDR = 4.17 *×* 10*^−^*^2^), myofibril assembly (FE*>* 100, FDR = 2.98 *×* 10*^−^*^2^), cardiac muscle contraction (FE*>* 100, FDR = 2.47 *×* 10*^−^*^2^), and regulation of striated muscle contraction (FE*>* 100, FDR = 2.87 *×* 10*^−^*^2^). Thus, although both target genes exhibit the specified enriched functions, an egocentric network analysis reveals that they do not share mutual genes with strong co-expression across the heart, artery, kidney, and skeletal muscle tissues. Therefore, while they may have functional overlap, their co-expressing gene partners diverge for each target gene.

In our comparative analysis of gene sets uniquely associated with either single or recurrent AF, we also evaluated genes that were consistent across both AF categories. Among the 16 genes identified for both AF groups (labelled as *Common* in Tab. 2), there was a significant up-regulation in tissues of the heart’s left ventricle, atrial appendage, and skeletal muscles (see Additional File 1, Fig. S11). Notably, these genes did not show any enrichment in specific gene ontology categories. Upon conducting an egocentric network analysis, we found that 14 out of these 16 genes displayed strong co-expression patterns with genes differentially expressed in the heart, arteries, kidneys, and skeletal muscles. Moreover, 13 of these genes were interconnected via 61 shared neighbor genes (see Fig.4 B) and Additional File 3). The 61 genes found in common for the 13 target genes show significant enrichment for fumarate metabolic process (FE *>* 100, FDR = 1.83 *×* 10*^−^*^2^), regulation of atrial cardiac muscle cell action potential (FE *>* 100, FDR = 2.51 *×* 10*^−^*^2^), regulation of mitochondrial RNA catabolic process (FE*>* 100, FDR = 3.36 *×* 10*^−^*^2^), mitochondrial acetyl-CoA biosynthetic process from pyruvate (FE *>* 100, FDR = 3.32 *×* 10*^−^*^2^), succinyl-CoA catabolic process (FE *>* 100, FDR = 4.10 *×* 10*^−^*^2^), tricarboxylic acid cycle (FE = 73.80, FDR = 5.34 *×* 10*^−^*^8^), branched-chain amino acid catabolic process (FE = 46.69, FDR = 1.11 *×* 10*^−^*^2^), inner mitochondrial membrane organization (FE= 35.33, FDR = 2.16 *×* 10*^−^*^3^), aerobic electron transport chain (FE = 15.20, FDR = 2.98 *×* 10*^−^*^2^), mitochondrial ATP synthesis coupled electron transport (FE = 14.37, FDR = 3.50 *×* 10*^−^*^2^), and alpha-amino acid catabolic process (FE = 14.21, FDR = 3.53 *×* 10*^−^*^2^).

Thus, the gene sets identified in common and uniquely for single and recurrent AF events are up-regulated in atrial appendage tissues of the heart, left ventricular tissues of the heart, and muscle skeletal tissues, where the gene set identified for recurrent AF and single AF events show significant up-regulation in atrial appendage and left ventricular tissues of the heart, respectively, while the gene set identified for both single and recurrent AF shows significant up-regulation of all three tissues. While both gene sets specifically identified for single and recurrent AF events show enrichment for functions related to heart and cardiac muscle, the recurrent AF gene set shows interesting enrichment of functions related to regulation of heart rate and cardiac muscle cell action potential. Collectively, these findings underscore the distinct genetic underpinnings between patients experiencing isolated versus recurrent AF events, with the recurrent AF genes revealing more intricate processes.

#### 3.4.2 Network MI

The 24 nearest genes to the regions specifically identified for single MI events (listed as Single in Tab. 3) show no significant enrichment for any gene ontology terms. Differential gene expression analysis shows no significant up or down regulation of these genes in any tissue, but significant expression levels in tibial nerve and high expression levels in tibial artery and aorta arty tissues (see Additional File 1, Fig S12).

When inspecting gene co-expression in heart, artery, kidney, and muscle skeletal tissues, we find that 21 of these genes show high co-expression with other genes in these tissues. Again creating egocentric networks for each of these 21 target genes, we find that all 21 target genes cluster in a giant component, where 110 genes are connected to two or more of them. The network depicted in Fig. 5 A) illustrates genes that are interconnected with multiple single MI target genes. Notably, there’s a dense cluster in the upper left portion of the figure, dominated by GLI2, GLIS3, TRIB3, and SFRP1, all of which are interconnected and share a majority of their neighborhood genes. The known associations of both GLI2 and SFRP1 with CVD-related traits, coupled with the detection of TRIB3 and SFRP1 in the comprehensive MI GWAS, suggest potential shared functional roles of these four genes. Furthermore, they also exhibit significant interconnections with BBS9, HECTD4, ATXN2, and SYNDIG1. It’s worth noting that HECTD4 and ATXN2 have recognized associations with MI.

**Figure 5.**
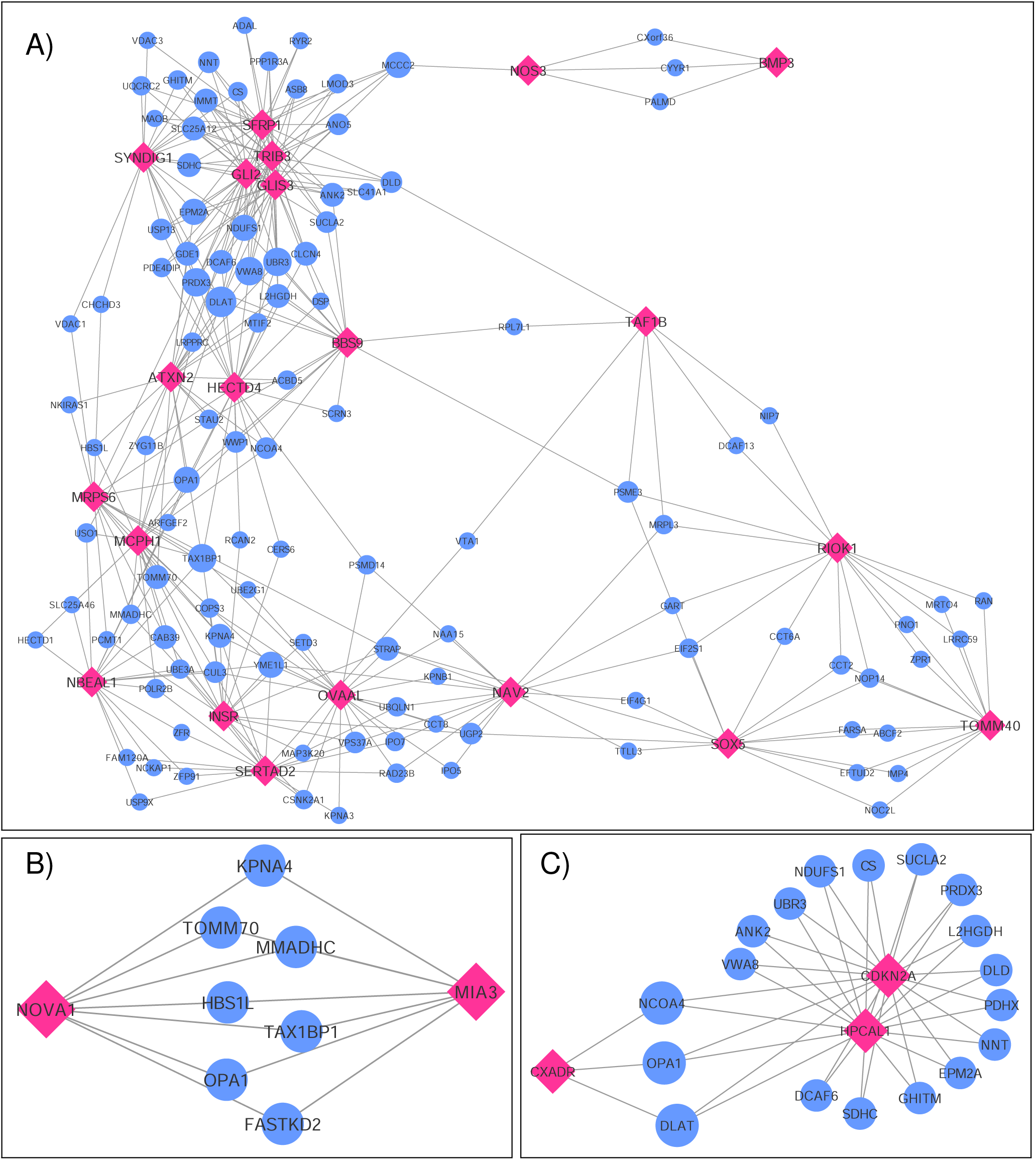
Networks showing the strongest shared neighbourhood of co-expressed genes for the GWAS (target) genes associated with A) single MI uniquely, B) recurrent MI uniquely and C) both single and recurrent MI. Pink diamond nodes represent the target genes and blue circular nodes represents the neighbouring genes. The sizes of the blue nodes are scaled according to their number of nearest neighbours in the network.

Gene ontology analysis of the 110 genes shared between two or more target genes (see Additional File 3) show significant enrichment for positive regulation of several functional categories, more specifically establishment of protein localization to telomere (*FE* = 54.18*, FDR* = 1.95 *×* 10*^−^*^2^), positive regulation of protein localization to Cajal body (*FE* = 49.26*, FDR* = 2.28 *×* 10*^−^*^2^), NLS-bearing protein import into nucleus (*FE* = 38.02*, FDR* = 3.85 *×* 10*^−^*^3^), positive regulation of telomerase RNA localization to Cajal body (*FE* = 36.12*, FDR* = 4.17 *×* 10*^−^*^2^), cristae formation (*FE* = 36.12, 4.00 *×* 10*^−^*^2^), tricarboxylic acid cycle (*FE* = 29.13*, FDR* = 1.59 *×* 10*^−^*^3^), regulation of proteasomal protein catabolic process (*FE* = 8.38*, FDR* = 1.63 *×* 10*^−^*^3^), regulation of ubiquitin-dependent protein catabolic process (*FE* = 7.35*, FDR* = 2.27 *×* 10*^−^*^2^), proteasome-mediated ubiquitin-dependent protein catabolic process (*FE* = 4.68*, FDR* = 4.76*×*10*^−^*^2^), purine-containing compound metabolic process (*FE* = 4.21*, FDR* = 4.69 *×* 10*^−^*^2^), nucleotide metabolic process (*FE* = 3.84*, FDR* = 4.69 *×* 10*^−^*^2^), and cellular component biogenesis (*FE* = 2.06*, FDR* = 3.57 *×* 10*^−^*^2^).

Upon analyzing the two genes uniquely associated with recurrent MI (listed as Recurrent in Tab. 3), we observe neither significant enrichment in gene ontology terms nor any distinctive expression patterns across tissue types (see Additional File 1, Fig. S13). In constructing egocentric networks for these genes, we identify seven overlapping genes (see Additional File 3) among the top-25 most correlated for each gene, as depicted in Fig. 5 B). While four of these seven neighboring genes (TOMM70, FASTKD2, MMADHC and OPA1) are related to mitochondrial function and energy production, gene ontology analysis yielded no significant enrichment.

Investigating the genes that are in common for both MI groups (listed as Common in Tab. 3), we find no significant differential co-expression in any of the tissues (see Additional File 1, Fig. S14). No gene ontology terms were found to be significantly enriched for these four genes. The egocentric networks (Fig. 5 C)) revealed that three of them are linked through 18 shared neighborhood genes (see Additional File 3). The 18 genes connecting the three target genes show significant enrichment for mitochondrial acetyl-CoA biosynthetic process from pyruvate (FE *>* 100, FDR = 5.92 *×* 10*^−^*^3^), tricarboxylic acid cycle (FE *>* 100, FDR= 5.37 *×* 10*^−^*^7^), negative regulation of release of cytochrome c from mitochondria (FE *>* 100, FDR= 4.88 *×* 10*^−^*^2^), cell redox homeostasis (FE= 81.70, FDR= 3.38 *×* 10*^−^*^3^), aerobic electron transport chain (FE = 39.90, FDR= 2.07 *×* 10*^−^*^2^), mitochondrial ATP synthesis coupled electron transport (FE = 37.71, FDR = 2.38 *×* 10*^−^*^2^), reactive oxygen species metabolic process (FE = 32.68, FDR = 3.25 *×* 10*^−^*^2^), and mitochondrion organization (FE = 12.85, FDR = 1.15 *×* 10*^−^*^2^).

In summary, MI gene sets do not exhibit notable tissue specificity or functional enrichment. Yet, the gene sets linked to both common and single MI incidents share multiple genes within their co-expression network neighborhoods. These shared genes exhibit functional enrichment for several pertinent processes. Conversely, the shared gene neighborhood specifically related to recurrent MI does not present any functional enrichment. However, some of the genes are related to mitochondrial function and energy production, similar to several of the enriched functions for the shared neighbouring genes for common MI genes. Notably, the neighbouring genes of the single MI genes shows significant enrichment for several function not identified for the recurrent of common neighbouring gene sets. Only one of the biological processes identified in the neighborhood of single MI genes is also seen in the neighborhood of common genes. This suggests unique functions associated with genes specific to single MI incidents, shedding light on potential reasons why certain patients experience only one MI event.

## 4 DISCUSSION

The study demonstrates distinct clinical characteristics and genetic predispositions between patients who experience a single atrial fibrillation/myocardial infarction (AF/MI) event and those with recurrent events. Notably, the single AF and single MI patient groups represent relapse-free patients, where we have filtered the data to ensure they have only experienced one event, with a buffer of at least 5 years for AF and 7 years for MI since the episode and before the study either ended or the patient died.

Single AF incidents appear more influenced by lifestyle factors and age, with only two unique genetic regions identified. In contrast, patients with recurrent AF are typically younger at their first event and exhibit a stronger genetic basis, as evidenced by 18 unique genetic regions linked to recurrent AF. Among these, two regions are near the NAV2 and SCN10A genes, previously hypothesized, but unconfirmed (36, 35) (*N* = 42, 585 East Asian population, *N* = 660 German population), to affect recurrent AF. The region with the lowest *p*-values and also the largest amount of significant SNPs was found near the PITX2 gene for both the single and the recurrent AF groups. Though PITX2 is the most known gene associated with recurrent AF (10, 7, 1) (*N* = 295 Turkish population, *N* = 195 Caucasian population, *N* = 991 German and American population), our results give no indication of such an effect. This observation extends to six other genes (SOX5, CAV1, EPHX2, ITGA9, SLC8A1, and TBX5) (35, 10, 8) (*N* = 660 German population, *N* = 295 Turkish population, *N* = 42, 585 East Asian population, *N* = 486 Caucasian population), where variants near the SOX5, CAV1 and TBX5 genes are identified in both the single and recurrent AF group, and no variants near the EPHX2, ITGA9 and SLC8A1 genes are identified in any of the groups. PheWAS analysis of these unique single and recurrent AF SNPs further highlight distinct susceptibilities: SNPs associated with single AF correlate with 44 phenocodes across various categories, whereas recurrent AF SNPs are linked to 91 phenocodes, predominantly in the circulatory system category. Network analysis of gene sets revealed that 16 of the 18 genes associated with recurrent AF are connected through 82 shared, highly co-expressed neighboring genes. These recurrent AF genes, along with their neighboring genes, are involved in complex processes related to heart rate regulation and cardiac muscle cell action potential. In contrast, the two genes associated with single AF events are linked to heart and cardiac muscle processes but do not share highly co-expressed genes.

We also find distinct clinical and genetic differences between patients with single and recurrent MI. Unlike AF, recurrent MI is more associated with older age at first MI event, lifestyle factors, and age-related issues. This is despite the fact that the single MI group is adjusted for early death due to MI and/or related comorbidities, and this should not affect the results. The genetic predisposition seems stronger in single MI cases, with a total of 24 uniquely associated regions. In contrast, recurrent MI is only associated with two regions that did not share the association with single MI cases. Of the 24 genetic regions uniquely identified for single MI, 14 are previously known for MI or other CVD-related traits. The remaining 10 are novel for MI and have previously not been reported for other CVD-related traits. While most of these novel regions consist of single SNPs primarily identified in the HUNT population, their nearest genes code for functions similar to known MI regions. Looking into the allele frequencies in each population (Additional File 4), we see that these variants are rather common in HUNT and in the general European population (based on reporting from gnomAD (37)), while they are rare variants in the UKBB population and thereby not included in the META-analysis. Further studies are needed to investigate if these variants show similar effects in other European and non-European populations.

PheWAS analysis reinforces the genetic distinction between single and recurrent MI groups. The single MI group’s SNPs are linked to 128 phenotypes, predominantly in the circulatory system category, with additional associations in the endocrine/metabolic category and notable links to neurodegenerative disorders. In contrast, the SNPs related to recurrent MI correlate with 16 phenocodes, involving both circulatory and endocrine/metabolic categories, but the associations are not as pronounced as those in the single MI group. Our network analysis reveals distinct gene interactions for each group. Of the 24 genes uniquely associated with single MI, 21 share connections with 110 genes in the co-expression network. However, the two genes associated with recurrent MI have seven highly co-expressed genes. This indicates more extensive genetic interconnections in single MI cases. Interestingly, the shared neighbouring genes for single MI, and those common to both single and recurrent MI, show functional enrichment in several biological processes. In contrast, the shared neighboring genes for recurrent MI do not show significant functional enrichment. This disparity suggests distinct biological pathways involved in single versus recurrent MI events. Moreover, only one function is common between the shared genes for both single and recurrent MI, suggesting unique biological mechanisms specific to single MI events.

The results suggest a greater genetic influence in atrial fibrillation (AF) compared to myocardial infarction (MI), but several factors could affect this perception. Clinically, it’s expected that more genes would increase the risk of recurrent AF, a pattern observed in this study. In contrast, the findings for MI are opposite, potentially influenced by their higher ages: MI becomes less common in younger individuals, and in older populations, comorbidities often overshadow genetic risk factors. There could also be physiological reasons behind these observations. For instance, MI in younger individuals might more frequently result from genetic factors related to platelet aggregation or atherosclerosis, conditions that are generally more responsive to treatment. In older individuals, MI might be more associated with broader age-related issues, reducing the relative impact of genetics. The reporting of these conditions could also influence the results, with AF potentially being under-reported compared to MI, which itself is possibly over-reported. This discrepancy could explain why phenocodes related to neurodegenerative disorders and cerebral ischemia emerge as significant only in single MI cases in the PheWAS results. These findings might be influenced by diagnostic practices where MI is often recorded as a cause of death, even when other diseases are the actual cause, or in cases where individuals die before a recurrent event occurs. The latter has been adjusted for by excluding individuals with single events with less than 7 years between the event and the censoring date, being either death or the end of the study. Still, even with this adjustment, some single MI event cases might be censored out before a second event, possibly influencing the the case groups and thereby the results.

The observed differences in clinical characteristics for both diseases might be even more pronounced if the HUNT and UK Biobank (UKBB) studies had used similar questionnaire formats. In the HUNT study, participants involved in both HUNT2 and HUNT3 provided cholesterol and blood pressure measurements taken eleven years apart. We opted to use the measurements recorded closest to the first AF or MI event, as we believe these offer the most relevant information. However, for the UKBB participants, most were measured only once, eliminating the option to choose measurements nearest to the event. This discrepancy in data collection methodology could explain why significant differences in cholesterol and blood pressure measurements are observed between single and recurrent event groups in the HUNT population but not in the UKBB group. The lack of longitudinal data in the UKBB may obscure potential differences that are more apparent in the HUNT study due to its repeated measurements.

In any GWAS study, larger sample sizes, diverse ancestry representation, and result replication are crucial. The HUNT study, comprised solely of individuals of European ancestry, led us to select only European ancestry participants from the UKBB for consistency. However, for global applicability, conducting similar analyses across all ancestries is essential. Regarding sample sizes, while the combined participant pool of HUNT and UKBB exceeds 500, 000, the specific filtering and subgrouping in our study result in some case/control groups having fewer than 2, 000 individuals. This reduced size may limit our ability to achieve robust significant findings. The small sample sizes might explain the prevalence of significant singleton SNP hits (one SNP per region) found only in the HUNT study, with the same SNPs with too rare allele frequencies in the UKBB population. Increasing sample sizes or including an additional population would likely enhance the reliability of these findings, particularly since these variants are reported as common in the general European population. While increasing the sample sizes could verify/dispute or even identify additional regions, some genetic differences observed between recurrent and single cases might diminish with larger sample sizes. Although many spikes in the Manhattan plots (Fig.1 and Fig.2) indicate clear differences between single and recurrent cases, certain regions almost reach significance for the opposite group, such as the hit on chromosome 8 for recurrent AF. This suggests that some observed genetic distinctions might be less pronounced with a more substantial and diverse sample.

While our meta-analysis did not yield any GWAS significant results when directly comparing recurrent to single AF or MI cases, the data still indicate genetic differences between these groups. Notably, some genetic regions were identified in both single and recurrent groups (termed ‘common’), while a substantial number were unique to each group. This could suggest that the common regions may play a role in the general susceptibility to AF and MI, whereas the unique regions could confer specific genetic risks for the disease’s form (single or recurrent events). Further studies are needed to test the hypotheses generated from this study, in particular investigate the effect of the novel SNPs and genes identified and their involvement in either single or recurrent AF/MI. While many of the identified genes and their related function might not have an direct confirmed effect on AF/MI, we do see some similar functions of the novel genes as for the known AF/MI genes, showing the potential of identification and understanding of new genetic functions of the diseases.

There has been considerable research aimed at uncovering genetic causes for AF and MI. Our findings underscore the importance of genetic studies focused on disease subgroups, as conducted here. Both AF and MI are widespread diseases with varied impacts on individuals’ lives. The progression and outcomes of these diseases are not uniform across all patients. By examining subgroups within these diseases, we could gain new insights into their mechanisms, potentially leading to more effective prevention and treatment strategies tailored to different patient profiles.

## DATA AVAILABILITY STATEMENT

The Trøndelag Health Study (HUNT) has invited persons aged 13 *−* 100 years to four surveys between 1984 and 2019. Comprehensive data from more than 140, 000 persons having participated at least once and biological material from 78, 000 persons were collected. The data are stored in HUNT databank and the biological material in HUNT biobank. HUNT Research Centre has permission from the Norwegian Data Inspectorate to store and handle these data. The key identification in the data base is the personal identification number given to all Norwegians at birth or immigration, whilst de-identified data are sent to researchers upon approval of a research protocol by the Regional Ethical Committee and HUNT Research Centre. To protect participants’ privacy, HUNT Research Centre aims to limit storage of data outside HUNT databank, and cannot deposit data in open repositories. HUNT databank has precise information on all data exported to different projects and are able to reproduce these on request. There are no restrictions regarding data export given approval of applications to HUNT Research Centre. For more information see: http://www.ntnu.edu/hunt/data.

The UKBB characteristics and genetic data are available for all researchers upon requested. To identify the number of events per participant, the HESIN-tables were applied. See https://biobank.ndph.ox.ac.uk/showcase/ for further information.

PheWas analysis were conducted on the Pan UKBB resource (24). See https://pan.ukbb.broadinstitute.org. The mapping from ICD9 and ICD10 codes are available at https://phewascatalog.org/phecodes.

Gene expression matrices were downloaded from the GTEx portal (30), V8. See https://www.gtexportal.org.

## ETHICS STATEMENT

Participation in the HUNT Study is based on informed consent, and the study has been approved by the Norwegian Data Inspectorate and the Regional Ethics Committee for Medical Research in Norway (REK: 2014/144). All participants from both UKBB and HUNT who have contributed in this study have provided their written informed consent to participate.

## AUTHOR CONTRIBUTIONS

IS identified the research topic. MH and AHS conceived and designed the study, with that defining the traits and cohorts, planed the analysis and analysed the characteristics of the cohorts. MH conducted the analysis and interpreted the results. MH wrote the manuscript. MH, EA, AHS, IS and HD edited and revised the manuscript. MH, AHS, IS, HD and EA approved the final version of the manuscript.

## FUNDING

M.H. and E.A. thank the K. G. Jebsen Foundation for Grant SKGJ-MED-015.

## Data Availability

All data produced in the present study are available upon reasonable request to the authors

## ACKNOWLEDGMENTS

The Trøndelag Health Study (HUNT) is a collaboration between HUNT Research Centre (Faculty of Medicine and Health Sciences, Norwegian University of Science and Technology NTNU), Trøndelag County Council, Central Norway Regional Health Authority, and the Norwegian Institute of Public Health. The genetic investigations of the HUNT Study, is a collaboration between researchers from the K.G. Jebsen center for genetic epidemiology and University of Michigan Medical School and the University of Michigan School of Public Health. The K.G. Jebsen Center for Genetic Epidemiology is financed by Stiftelsen Kristian Gerhard Jebsen; Faculty of Medicine and Health Sciences, NTNU, Norway. We would like to thank clinicians and other employees at Nord-Trøndelag Hospital Trust for their support and for contributing to data collection in this research project. We would also like to thank all participants that have contributed in the UKBB study.

## CONFLICT OF INTEREST STATEMENT

The authors confirm that there are no conflicts of interest. The authors declare that the research was conducted in the absence of any commercial or financial relationships that could be construed as a potential conflict of interest.

## SUPPLEMENTAL DATA

Additional File 1 - Fig. S1-S14. S1: QQ-plot of single AF vs. healthy controls in HUNT. S2: QQ-plot of single AF vs. healthy controls in UKBB. S3: QQ-plot of recurrent AF vs. healthy controls in HUNT. S4: QQ-plot of recurrent AF vs. healthy controls in UKBB. S5: QQ-plot of single MI vs. healthy controls in HUNT. S6: QQ-plot of single MI vs. healthy controls in UKBB. S7: QQ-plot of recurrent MI vs. healthy controls in HUNT. S8: QQ-plot of recurrent MI vs. healthy controls in UKBB. S9: Tissue specificity of the 18 genes uniquely identified for recurrent AF. Test results in each row are Bonferroni corrected and red bars show tissues where the set of genes are significantly up-regulated (top), down-regulated (middle), and both-sided (bottom). S10: Tissue specificity for the two genes identified uniquely for single AF. Test results in each row are Bonferroni corrected and red bars show tissues where the set of genes are significantly up-regulated (top), down-regulated (middle), and both-sided (bottom). S11: Tissue specificity for the 16 genes identified for both single and recurrent AF. Test results in each row are Bonferroni corrected and red bars show tissues where the set of genes are significantly up-regulated (top), down-regulated (middle), and both-sided (bottom). S12: Tissue specificity for the 24 genes identified uniquely for single MI. Test results in each row are Bonferroni corrected and red bars show tissues where the set of genes are significantly up-regulated (top), down-regulated (middle), and both-sided (bottom). S13: Tissue specificity for the two genes identified uniquely for recurrent MI. Test results in each row are Bonferroni corrected and red bars show tissues where the set of genes are significantly up-regulated (top), down-regulated (middle), and both-sided (bottom). S14: Tissue specificity for the four genes identified for both single and recurrent MI. Test results in each row are Bonferroni corrected and red bars show tissues where the set of genes are significantly up-regulated (top), down-regulated (middle), and both-sided (bottom). S15-S39: Regional plot of single significant hit identified uniquely for recurrent AF, single MI or recurrent MI.

Additional File 2 - Excel file containing all phenocodes found to be significantly associated with the set of SNPs associated with both single and recurrent AF/MI or uniquely for one of them.

Additional File 3 - Excel file containing genes shared between the GWAS associated genes (target genes) in the network analysis for genes associated with both single and recurrent AF/MI or uniquely for one of them.

Additional File 4 - Excel file containing allele frequency of SNPs identified in only one of the populations (listed as for instance *−*? in Tab. 2 and 3). The columns named ‘AF HUNT’, ‘AF UKBB’ and ‘AF European’ shows the allele frequency in the HUNT population, the UKBB population and as reported in gnomAD (37)(https://gnomad.broadinstitute.org/), respectively. The ‘HUNT/UKBB’ columns shows if the SNP is identified in HUNT or UKBB, and the ‘imputation info’ columns shows the imputation info given in the genotyped UKBB files.

## Notes

### Competing Interest Statement

The authors have declared no competing interest.

### Funding Statement

This study was funded by K. G. Jebsen Foundation for Grant SKGJ-MED-015

### Author Declarations

The Norwegian Data Inspectorate and the Regional Ethics Committee for Medical Research in Norway (REK: 2014/144)

