## Additional File 1 for "Genome-wide association studies reveal differences in genetic susceptibility between single events versus recurrent events of atrial fibrillation and myocardial infarction: the HUNT study"

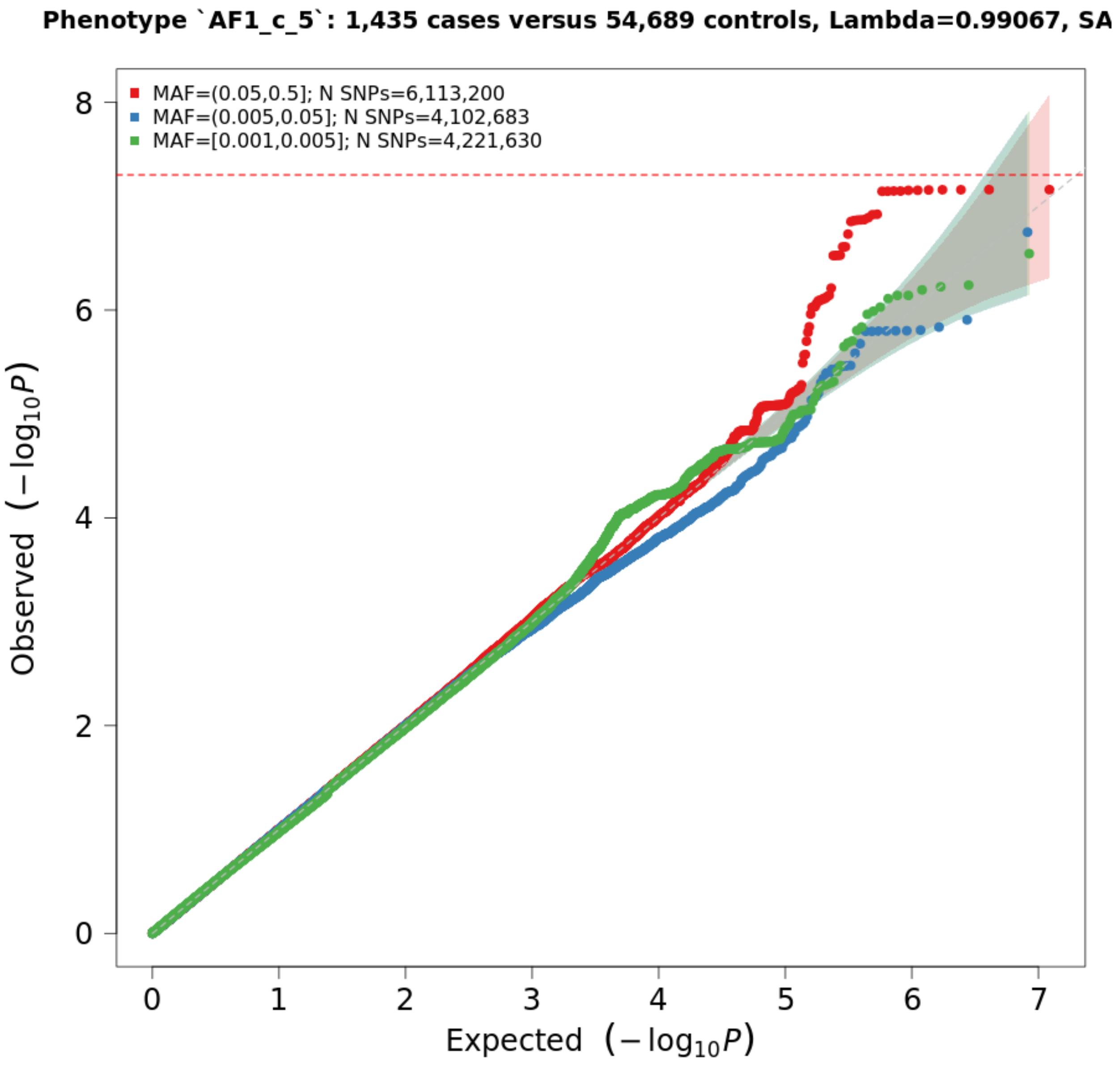


Fig. S1: QQ-plot of single AF vs. healthy controls in HUNT.

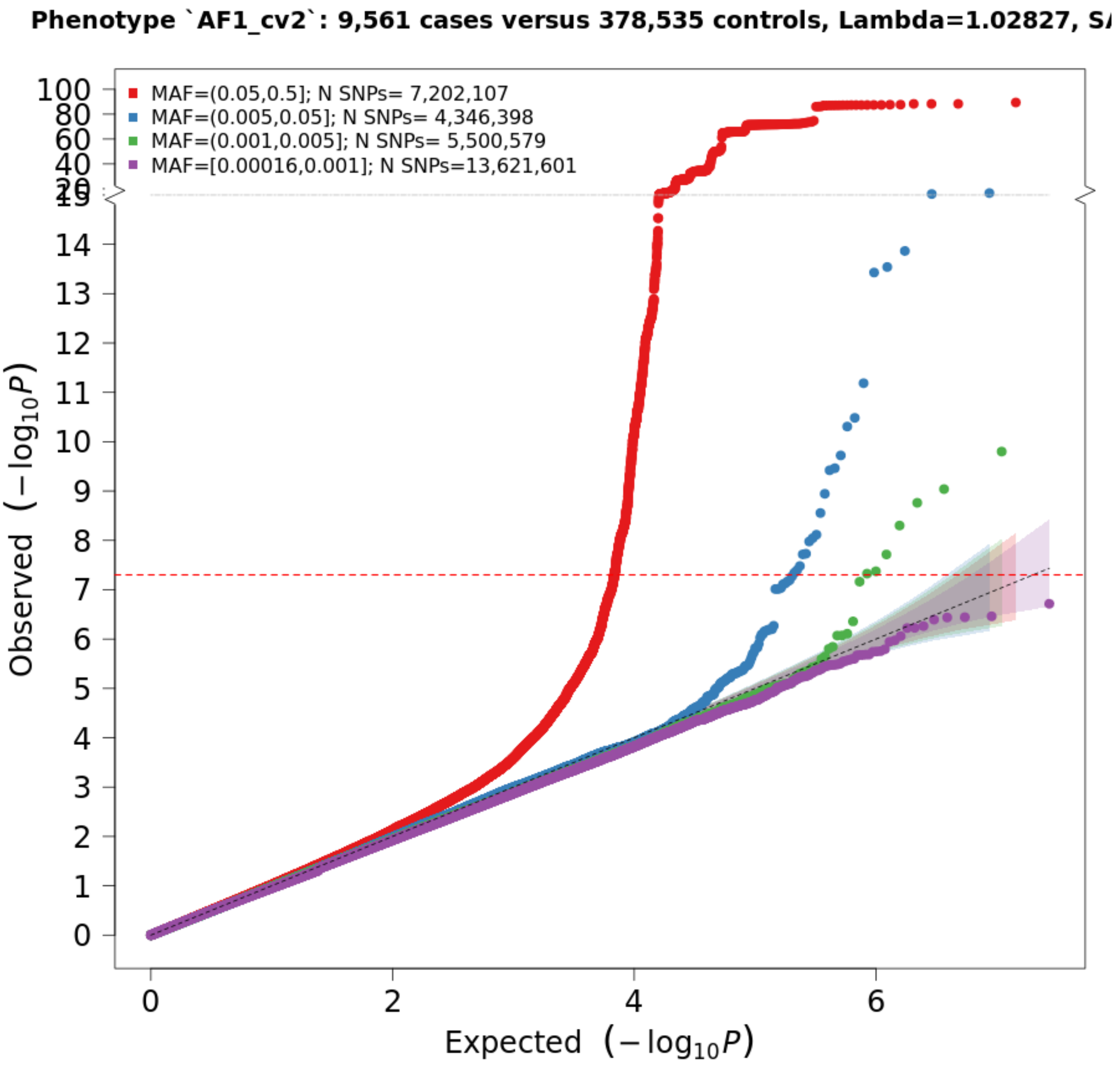


Fig. S2: QQ-plot of single AF vs. healthy controls in UKBB.

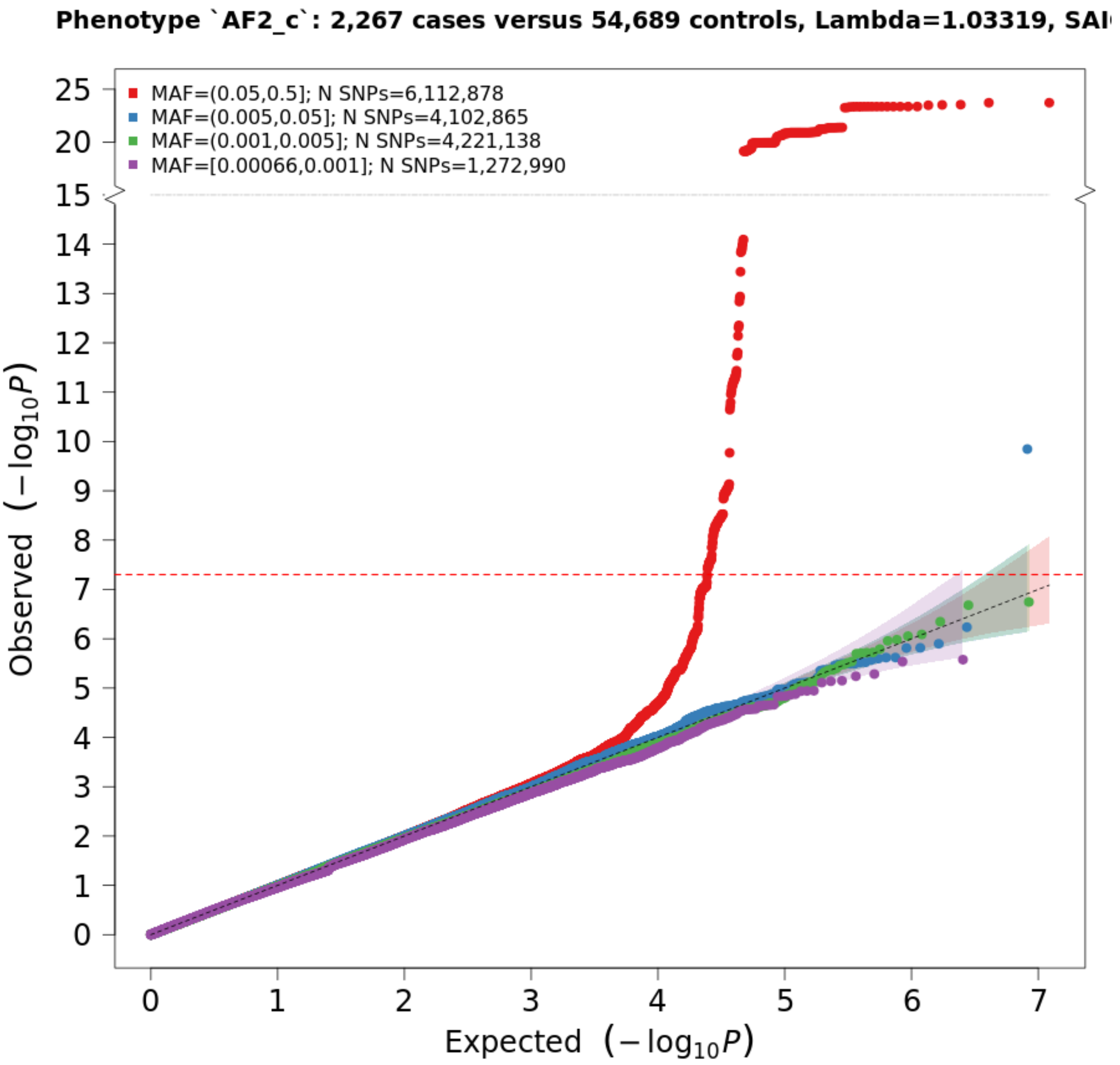


Fig. S3: QQ-plot of recurrent AF vs. healthy controls in HUNT.

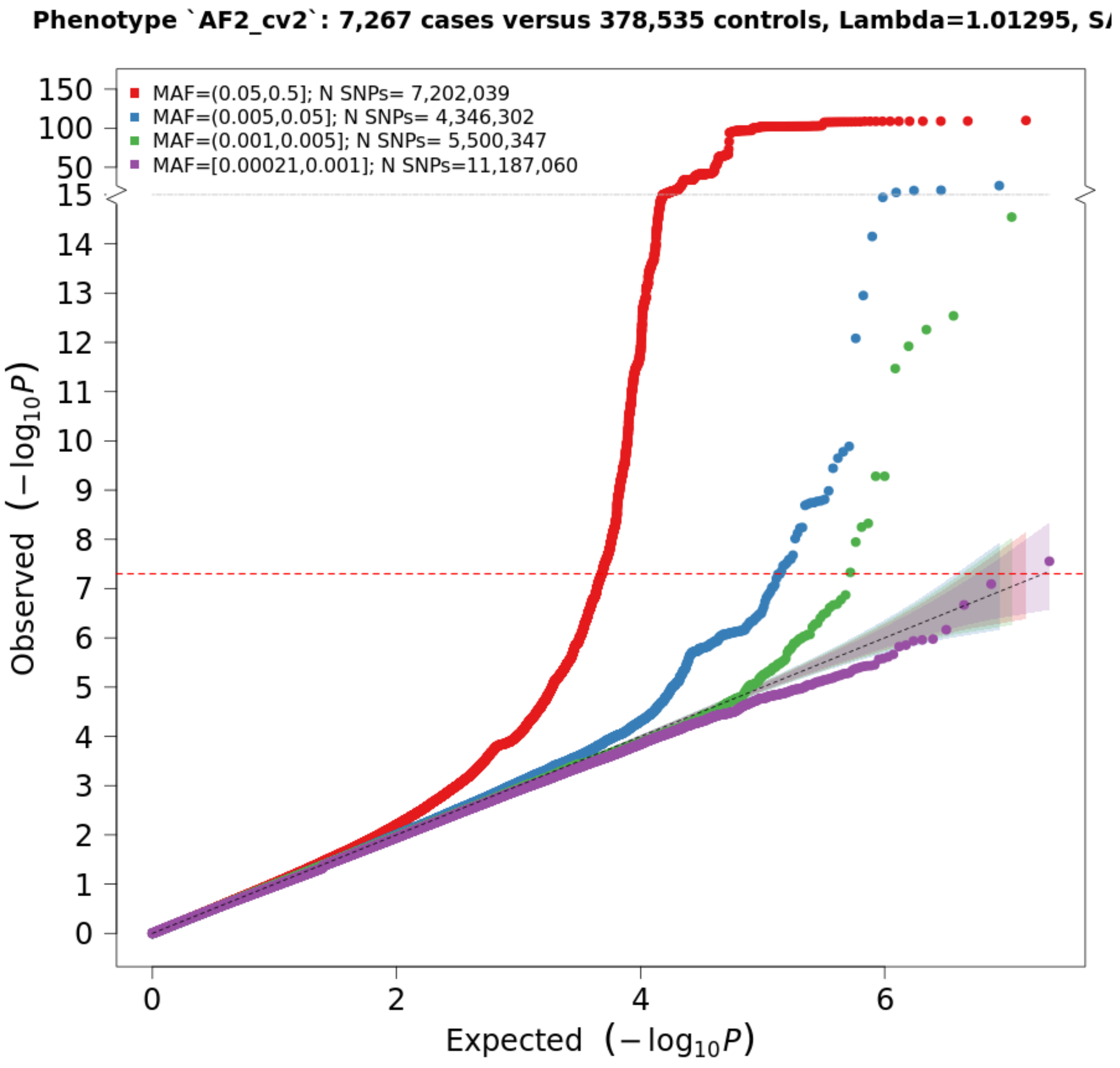


Fig. S4: QQ-plot of recurrent AF vs. healthy controls in UKBB.

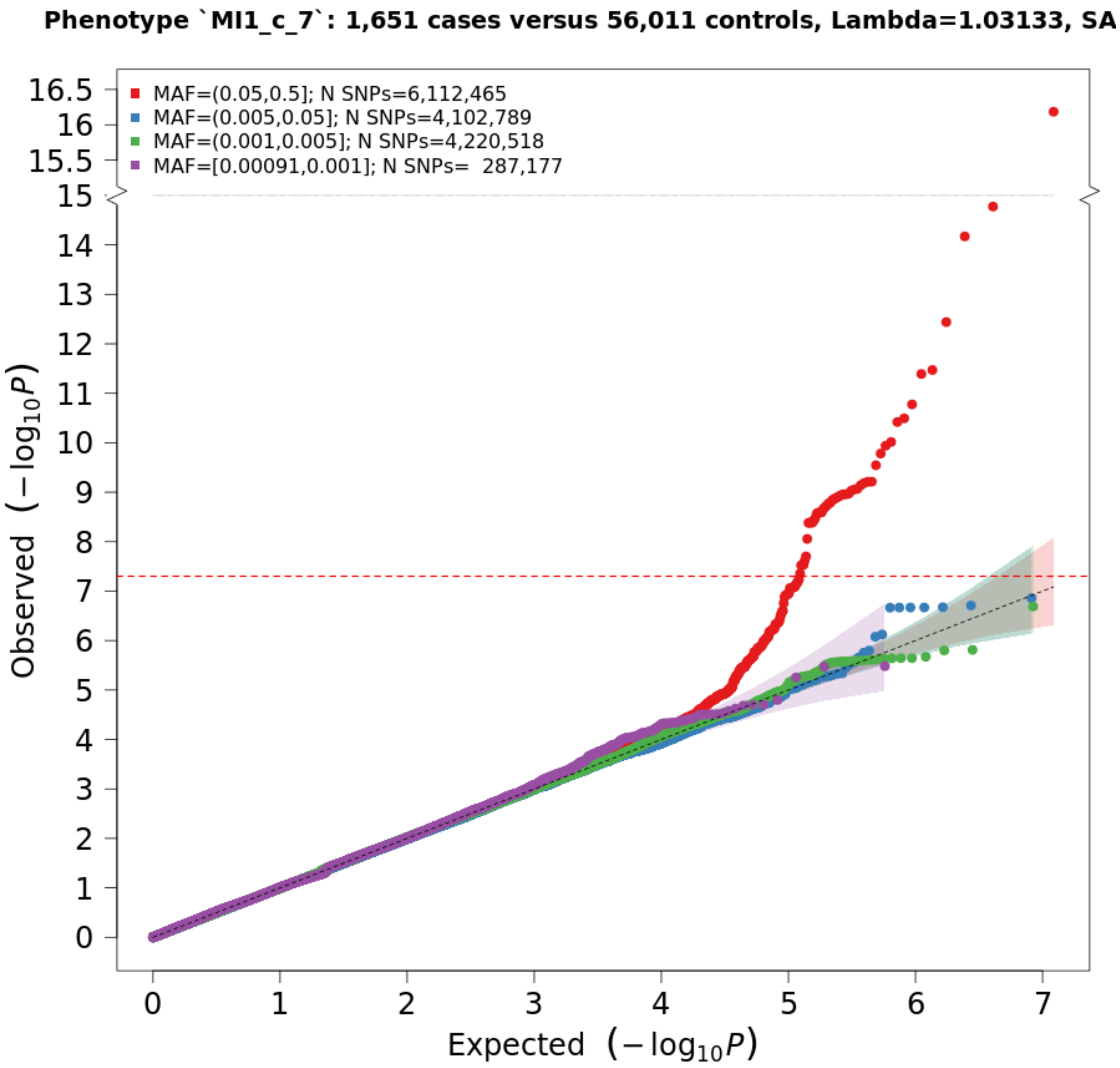


Fig. S5: QQ-plot of single MI vs. healthy controls in HUNT.

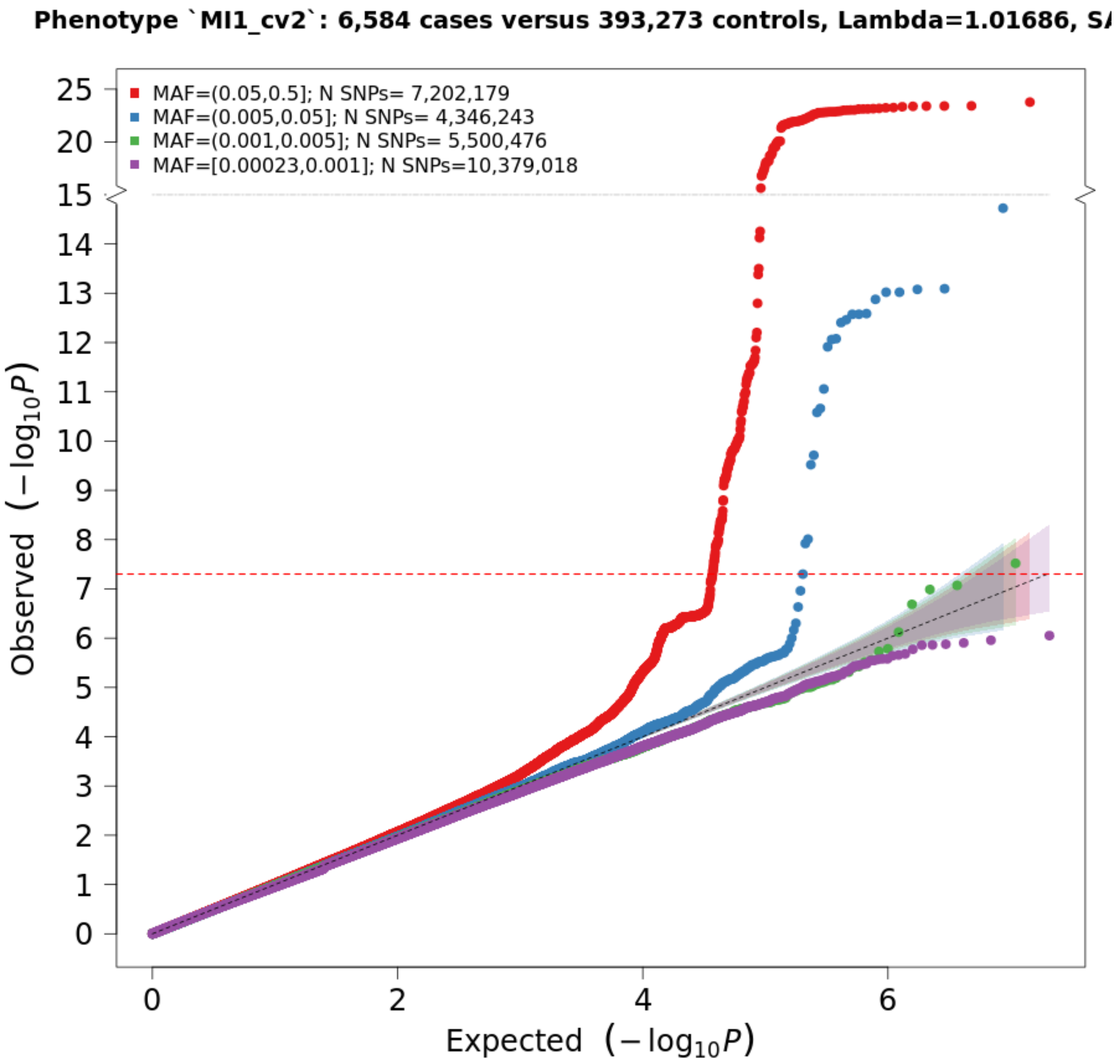


Fig. S6: QQ-plot of single MI vs. healthy controls in UKBB.

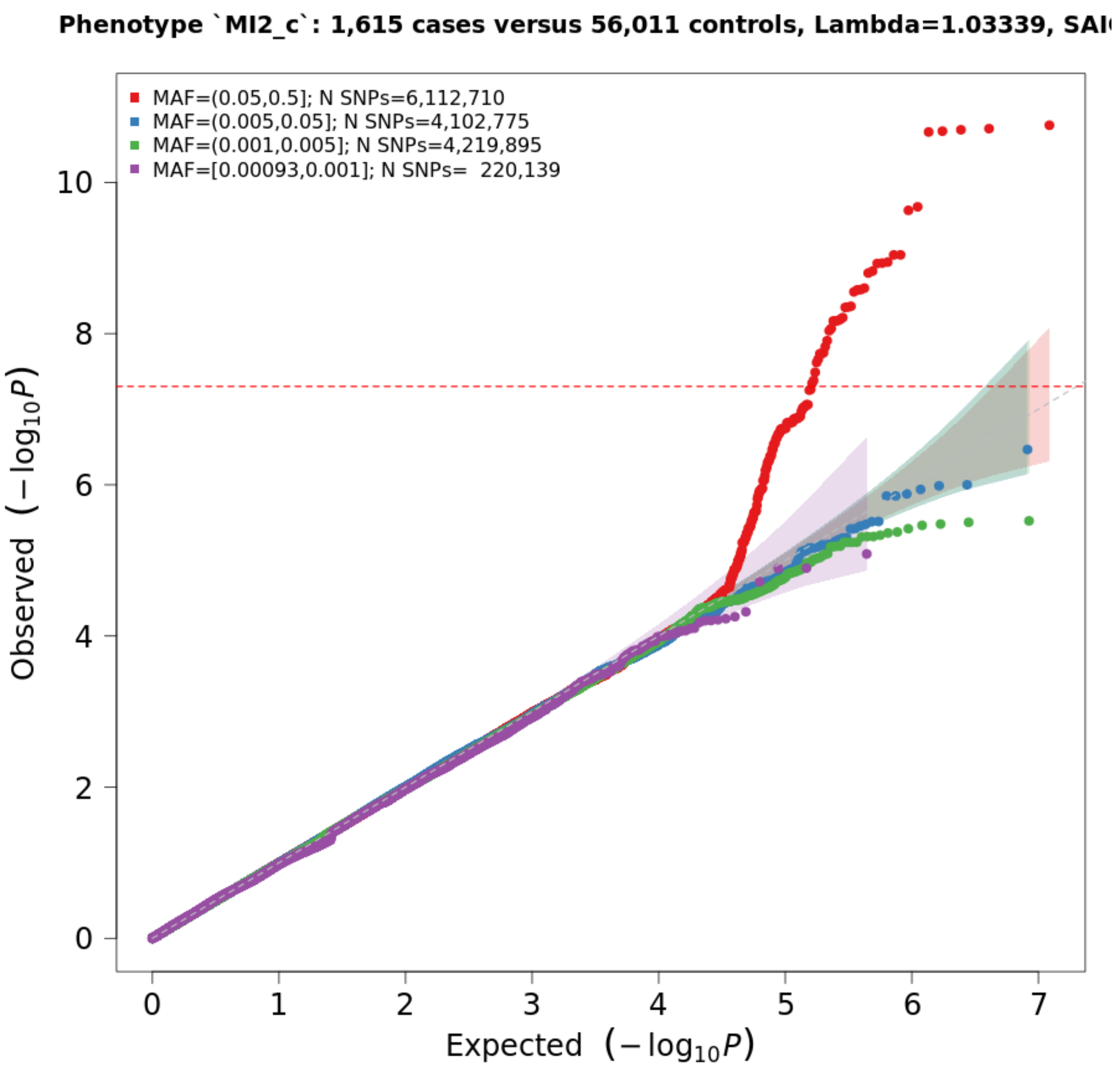


Fig. S7: QQ-plot of recurrent MI vs. healthy controls in HUNT.

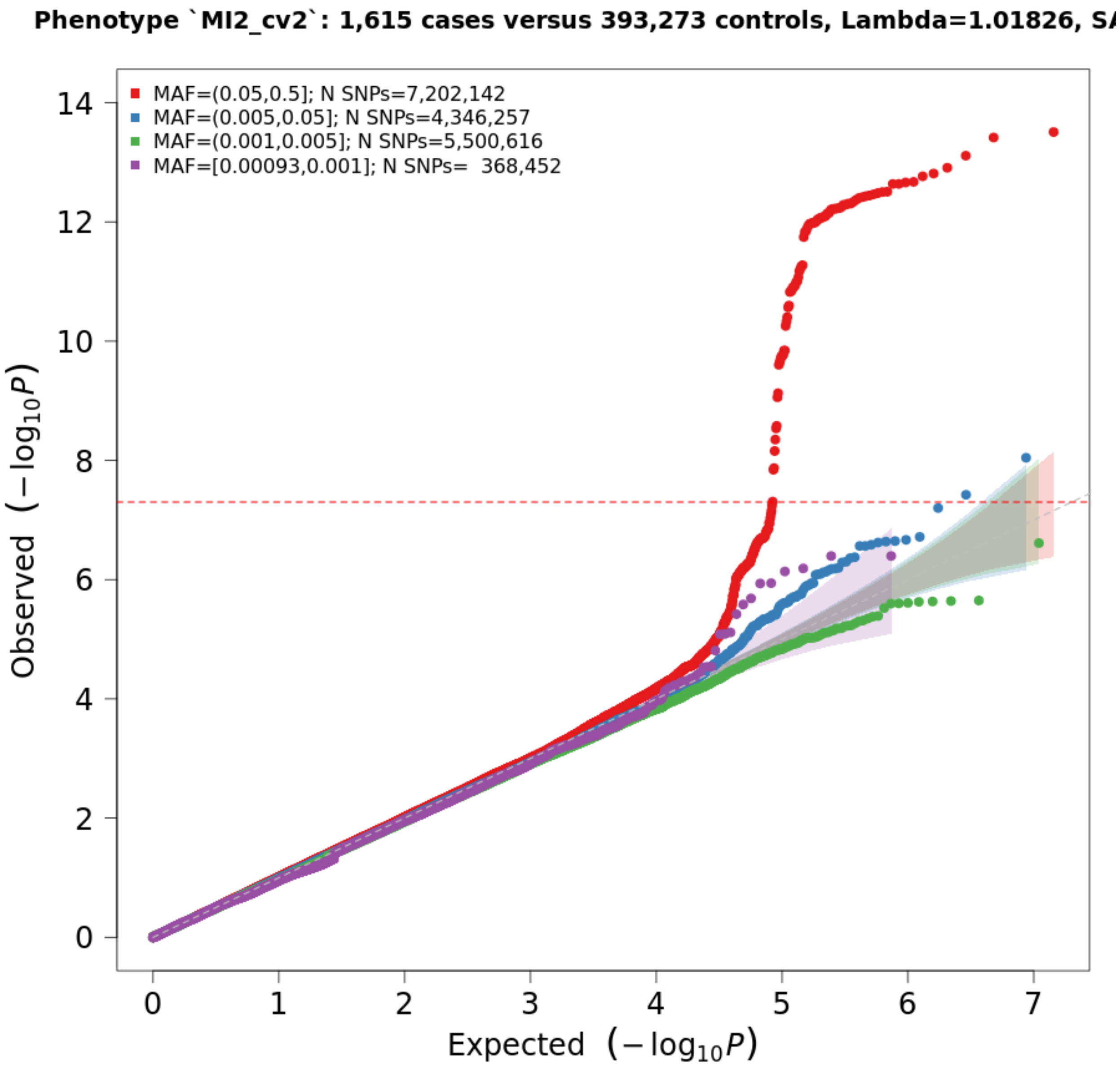


Fig. S8: QQ-plot of recurrent MI vs. healthy controls in UKBB.

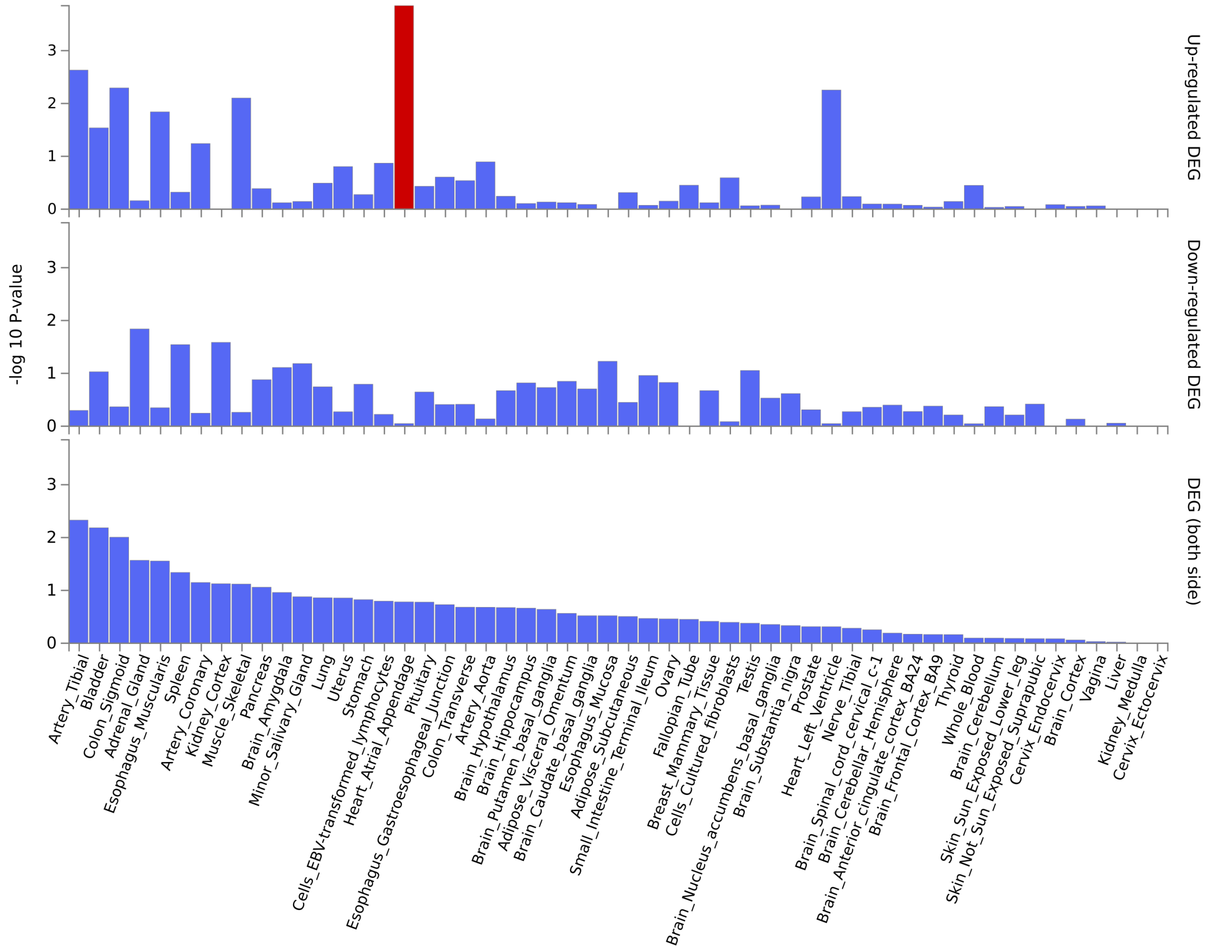


Fig. S9: Tissue specificity of the $18$ genes identified uniquely for recurrent AF. Test results in each row are Bonferroni corrected and red bars shows tissues where the set of genes are significantly up-regulated (top), down-regulated (middle) and both-sided (bottom).

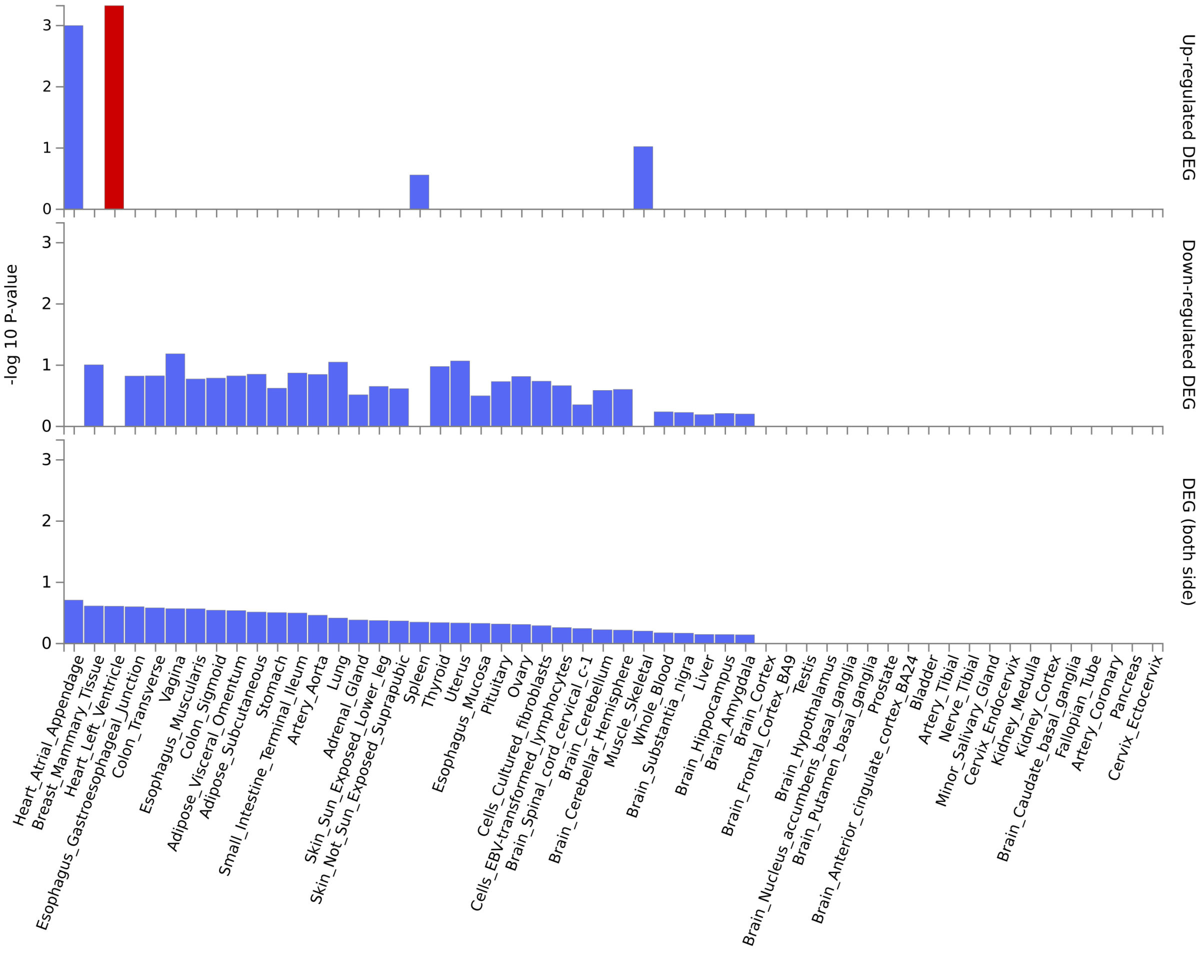


Fig. S10: Tissue specificity of the two genes identified uniquely for single AF. Test results in each row are Bonferroni corrected and red bars shows tissues where the set of genes are significantly up-regulated (top), down-regulated (middle) and both-sided (bottom).

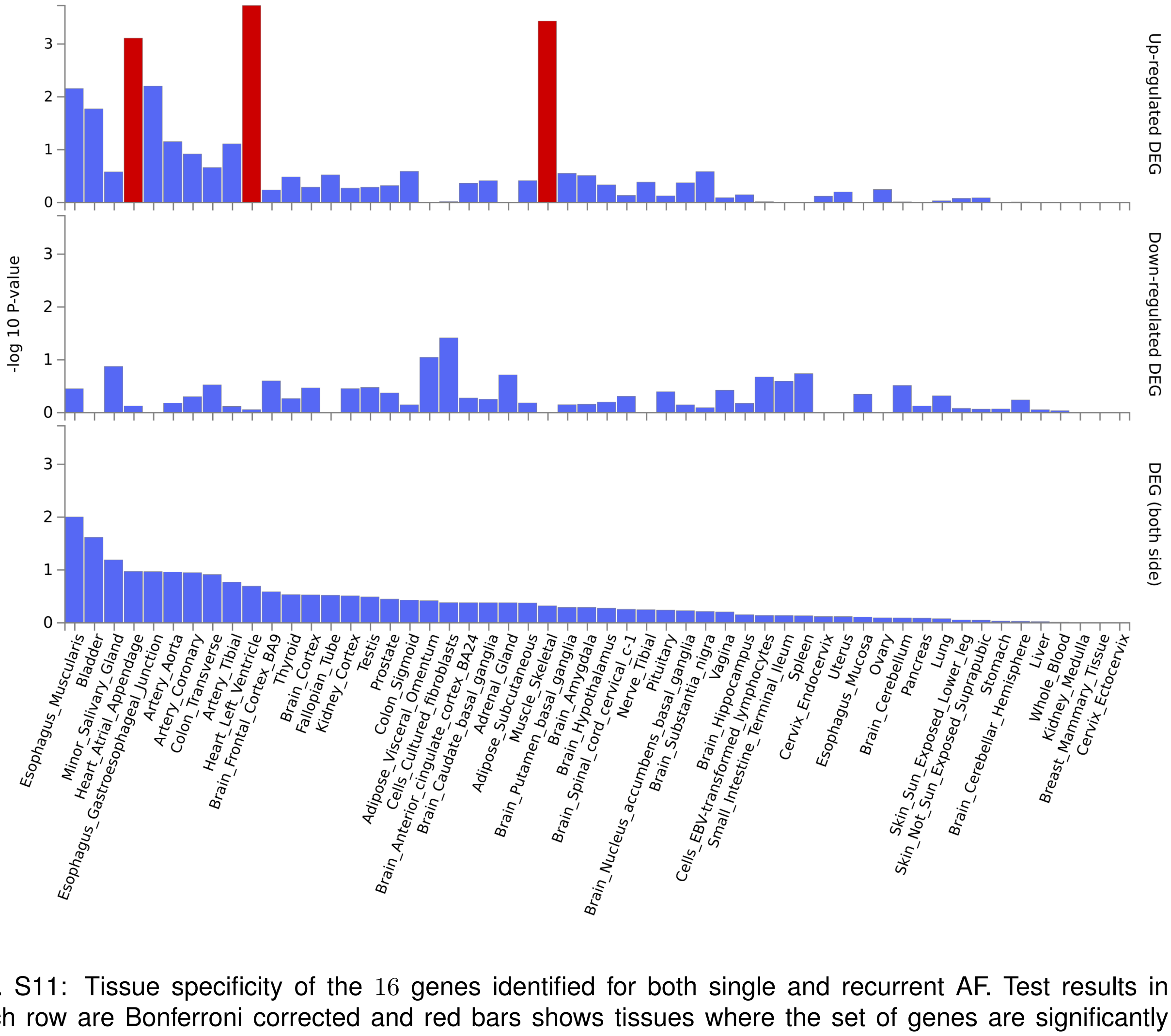


Fig. S11: Tissue specificity of the $16$ genes identified for both single and recurrent AF. Test results in each row are Bonferroni corrected and red bars shows tissues where the set of genes are significantly up-regulated (top), down-regulated (middle) and both-sided (bottom).

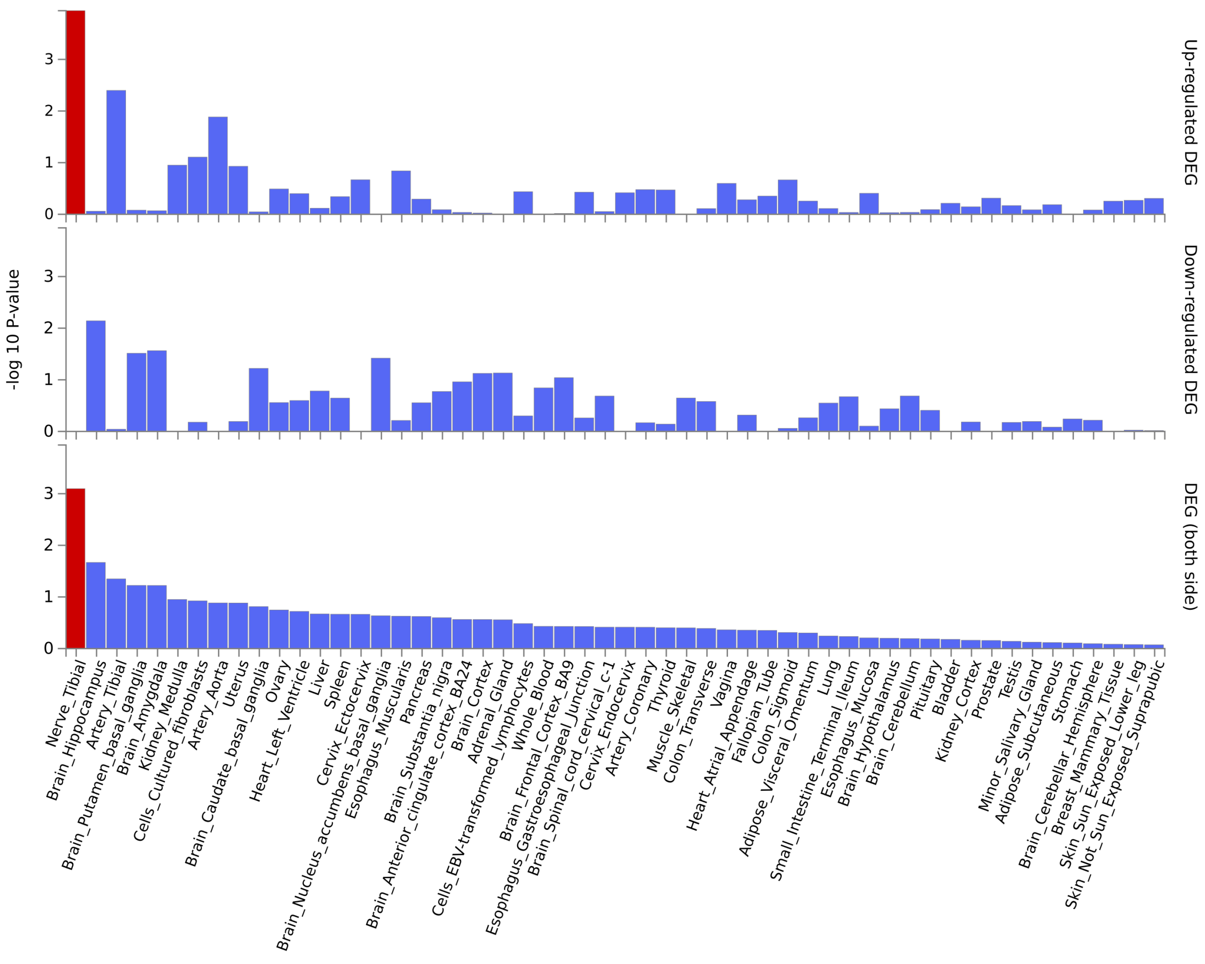


Fig. S12: Tissue specificity of the $24$ genes identified uniquely for single MI. Test results in each row are Bonferroni corrected and red bars shows tissues where the set of genes are significantly up-regulated (top), down-regulated (middle) and both-sided (bottom).

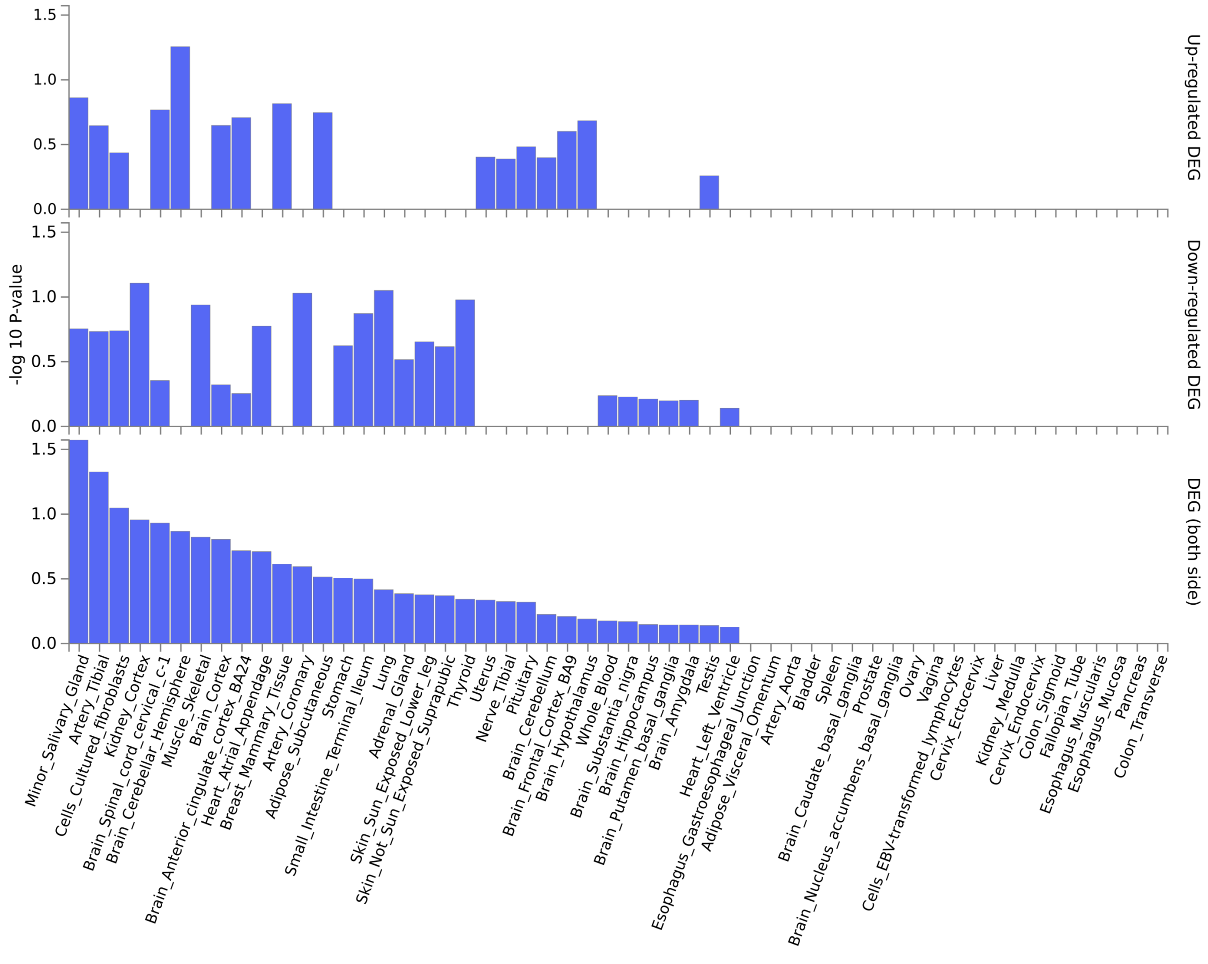


Fig. S13: Tissue specificity of the two genes identified uniquely for recurrent MI. Test results in each row are Bonferroni corrected and red bars shows tissues where the set of genes are significantly up-regulated (top), down-regulated (middle) and both-sided (bottom).

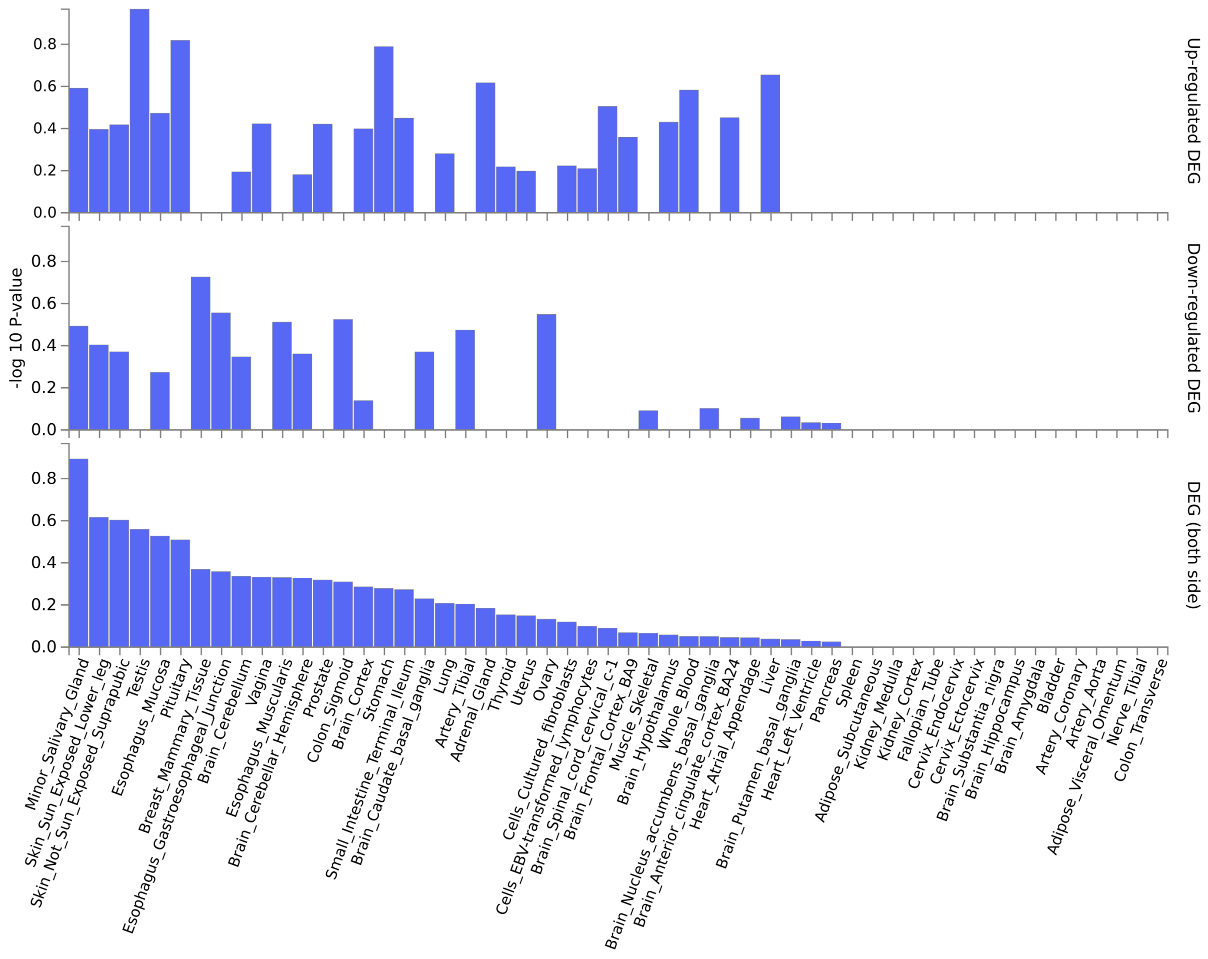


Fig. S14: Tissue specificity of the four genes identified for both single and recurrent MI. Test results in each row are Bonferroni corrected and red bars shows tissues where the set of genes are significantly up-regulated (top), down-regulated (middle) and both-sided (bottom).

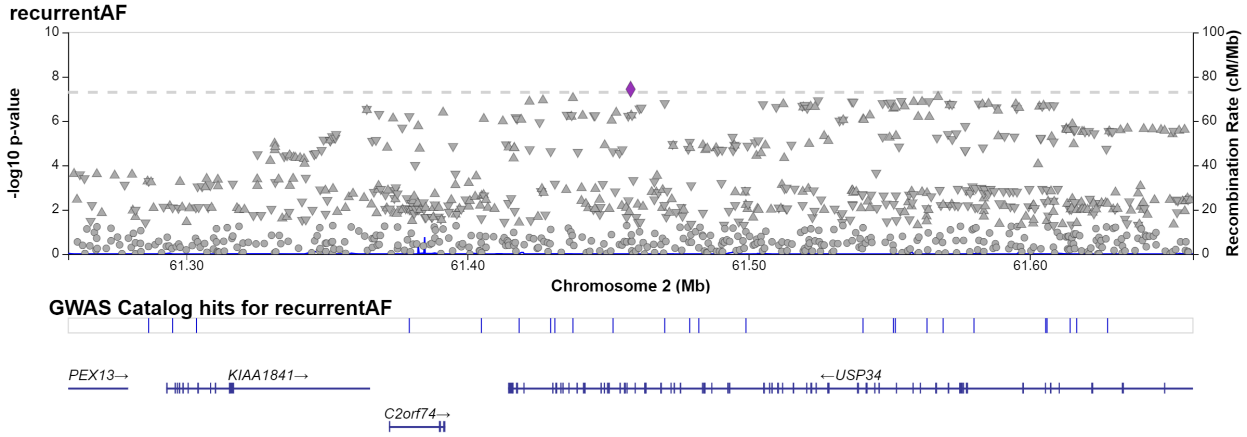


Fig. S15: Regional plot of single significant hit at chromosome 2 for recurrent AF.

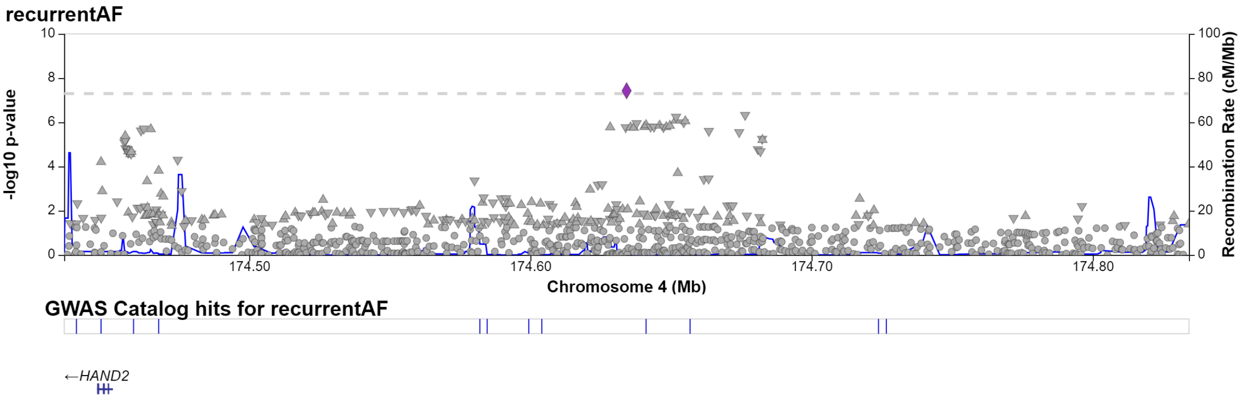


Fig. S16: Regional plot of single significant hit at chromosome 4 for recurrent AF.

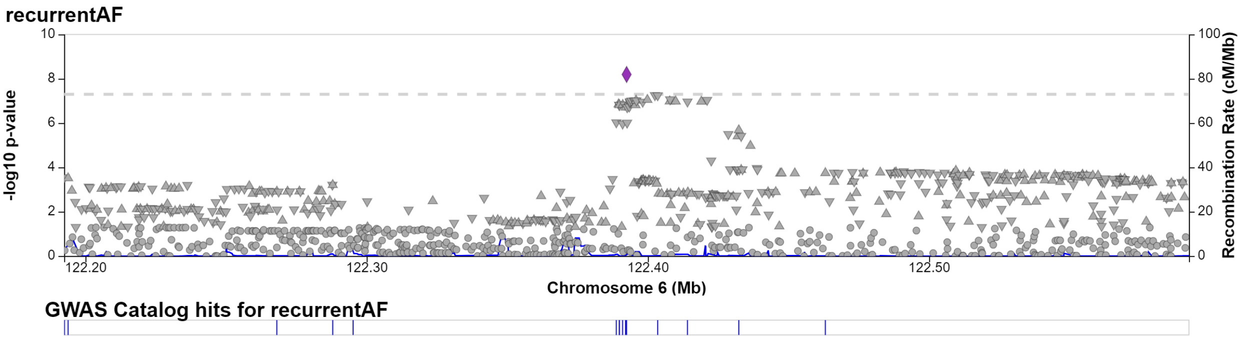


Fig. S17: Regional plot of single significant hit at chromosome 6 for recurrent AF.

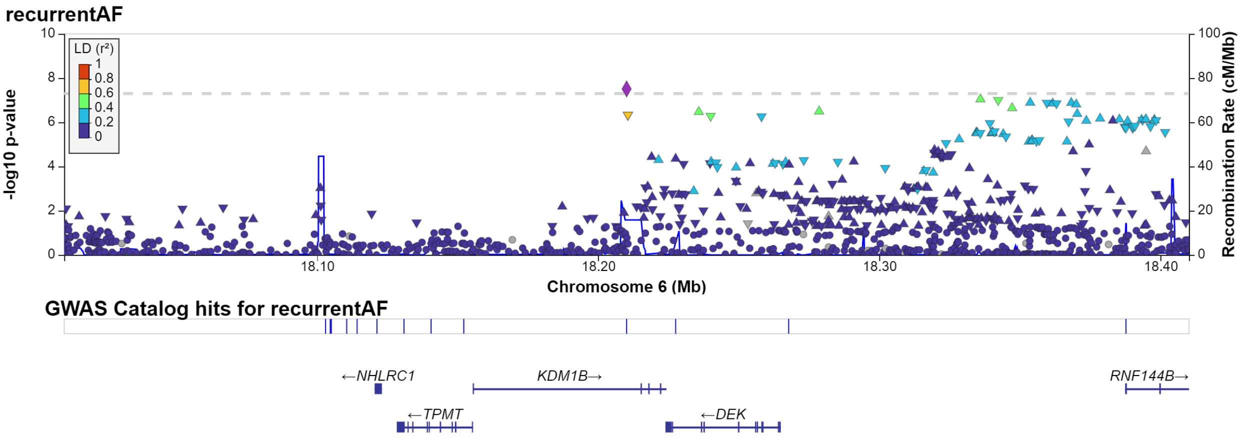


Fig. S18: Regional plot of single significant hit at chromosome 6 for recurrent AF.

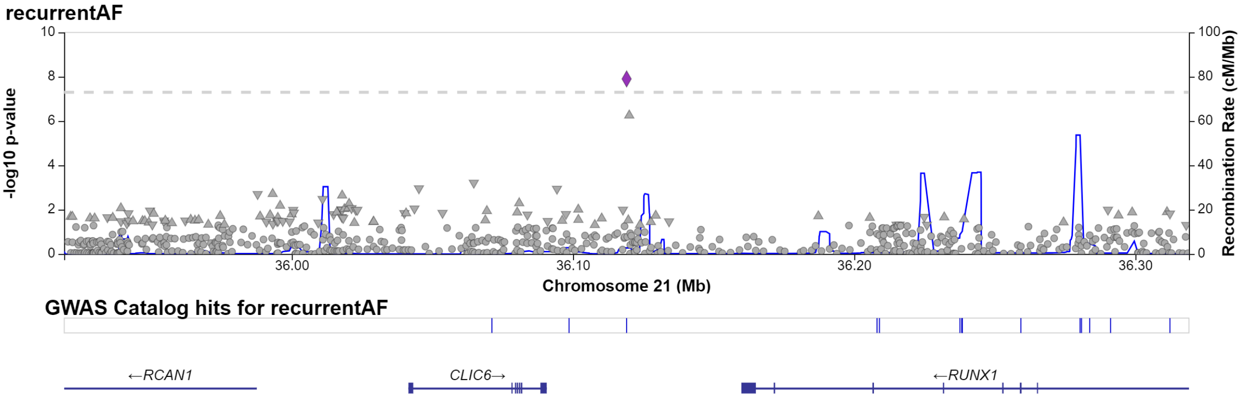


Fig. S19: Regional plot of single significant hit at chromosome 21 for recurrent AF.

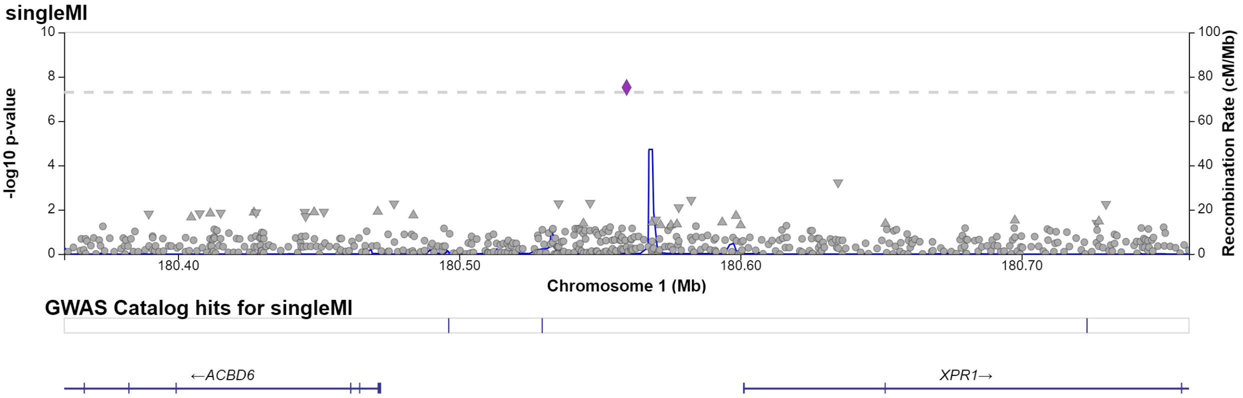


Fig. S20: Regional plot of single significant hit at chromosome 1 for single MI.

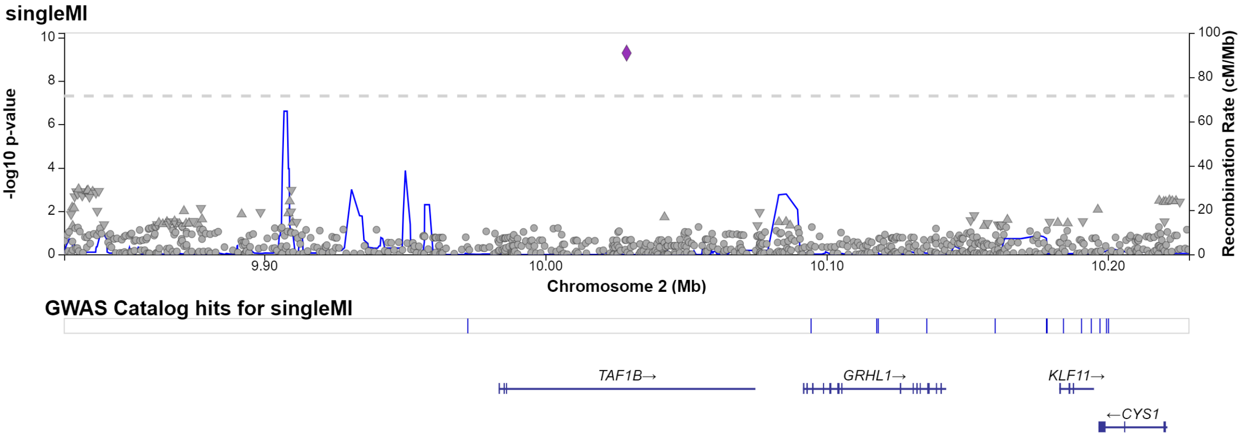


Fig. S21: Regional plot of single significant hit at chromosome 2 for single MI.

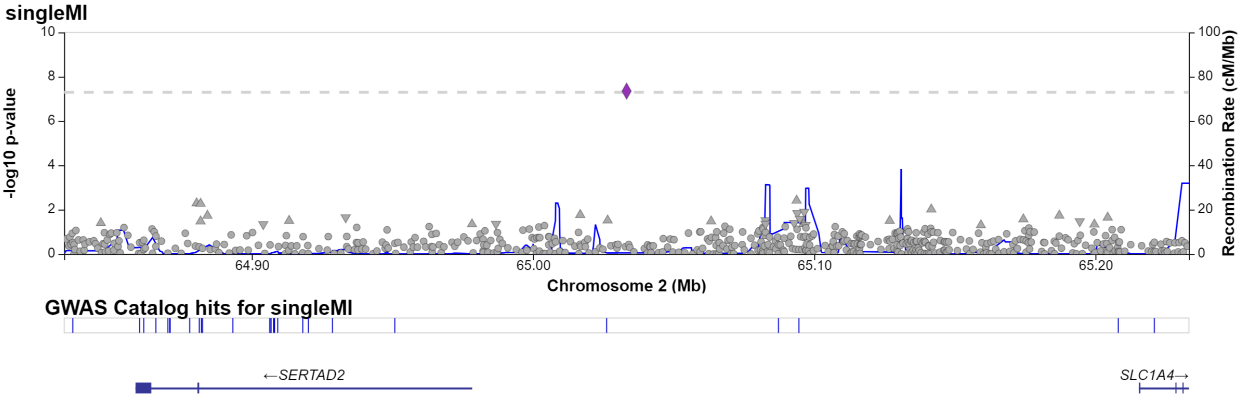


Fig. S22: Regional plot of single significant hit at chromosome 2 for single MI.

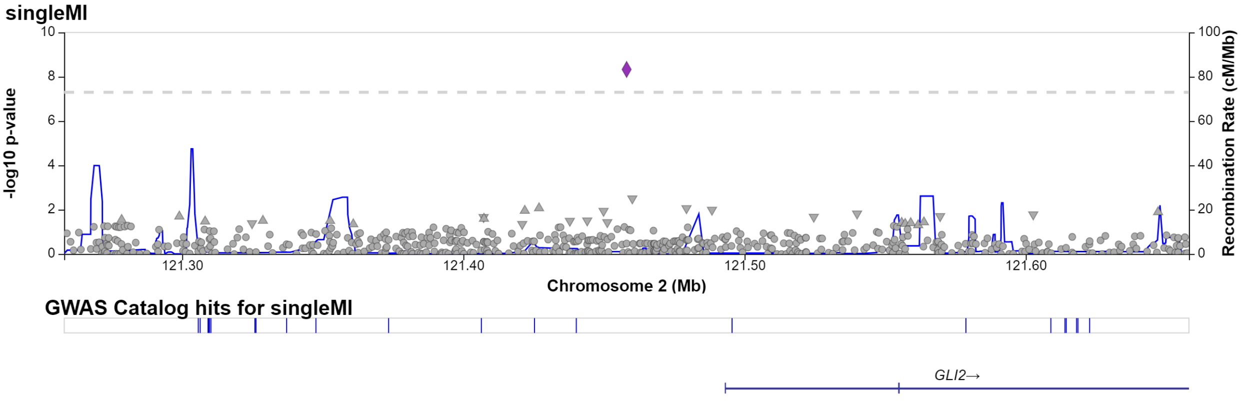


Fig. S23: Regional plot of single significant hit at chromosome 2 for single MI.

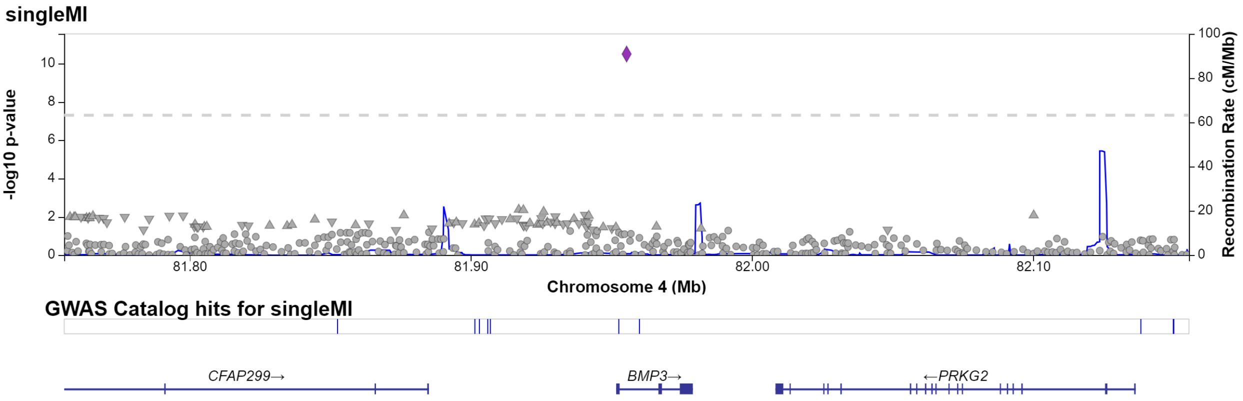


Fig. S24: Regional plot of single significant hit at chromosome 4 for single MI.

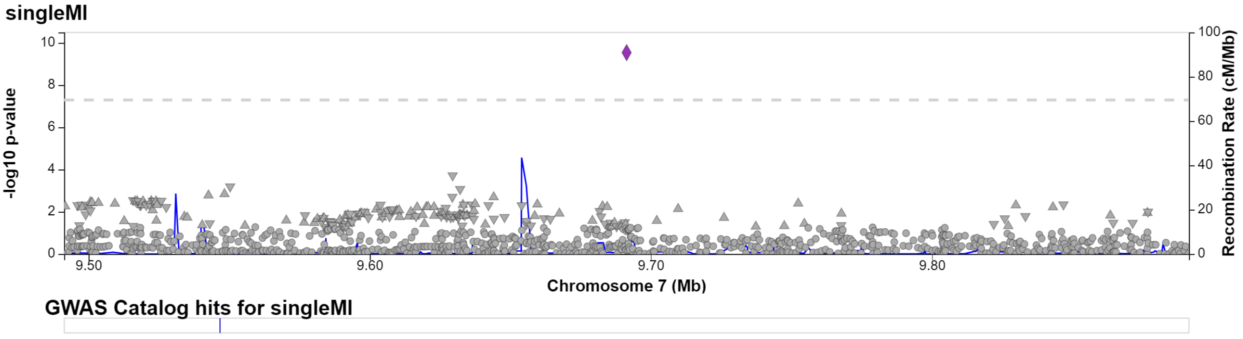


Fig. S25: Regional plot of single significant hit at chromosome 7 for single MI.

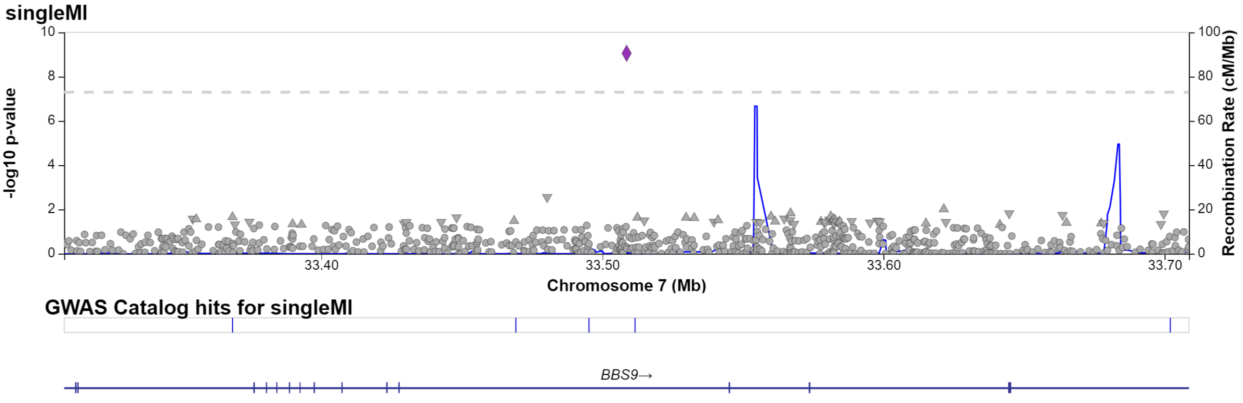


Fig. S26: Regional plot of single significant hit at chromosome 7 for single MI.

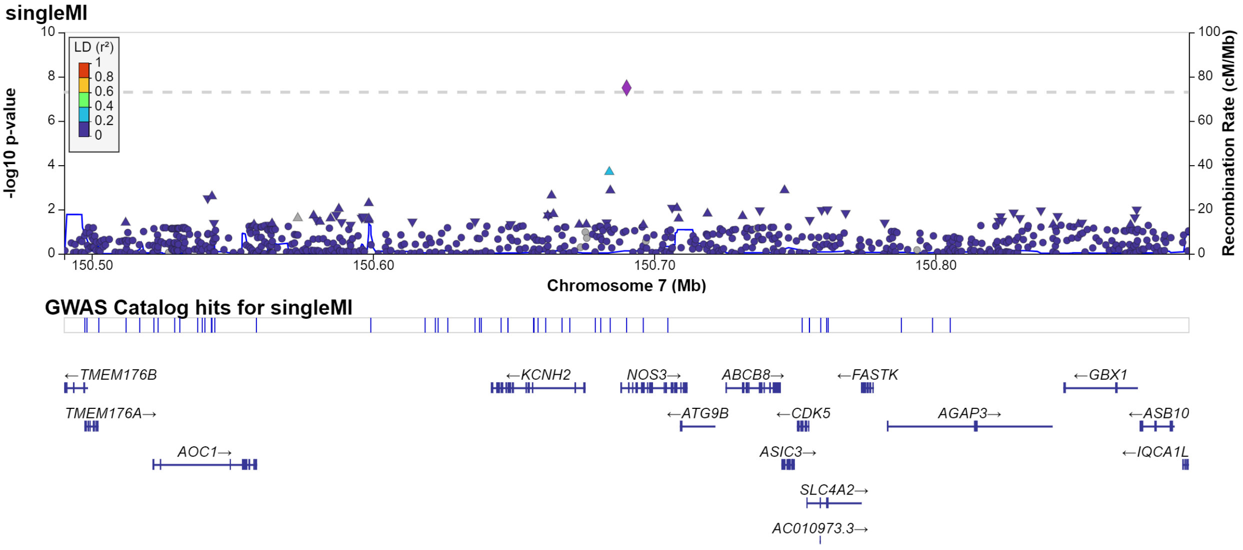


Fig. S27: Regional plot of single significant hit at chromosome 7 for single MI.

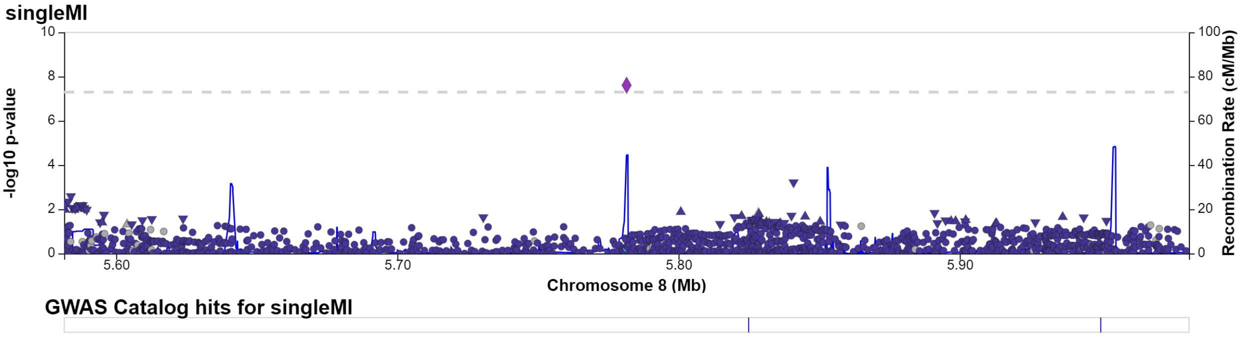


Fig. S28: Regional plot of single significant hit at chromosome 8 for single MI.

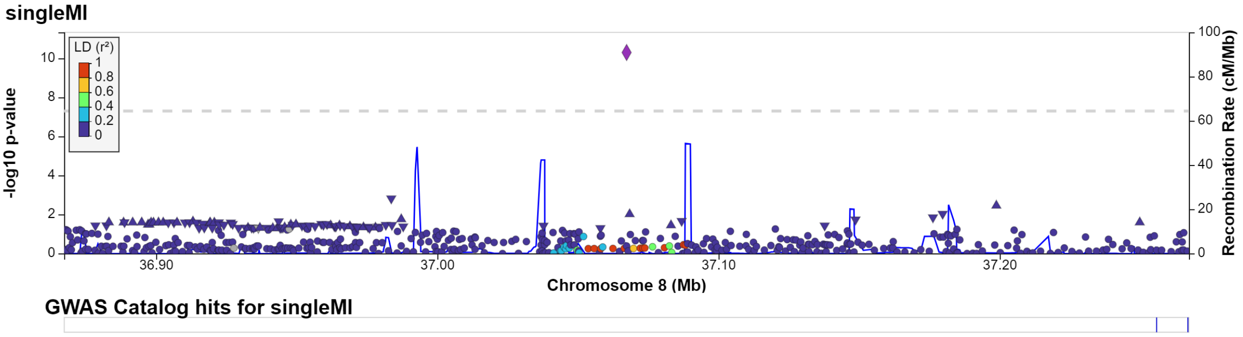


Fig. S29: Regional plot of single significant hit at chromosome 8 for single MI.

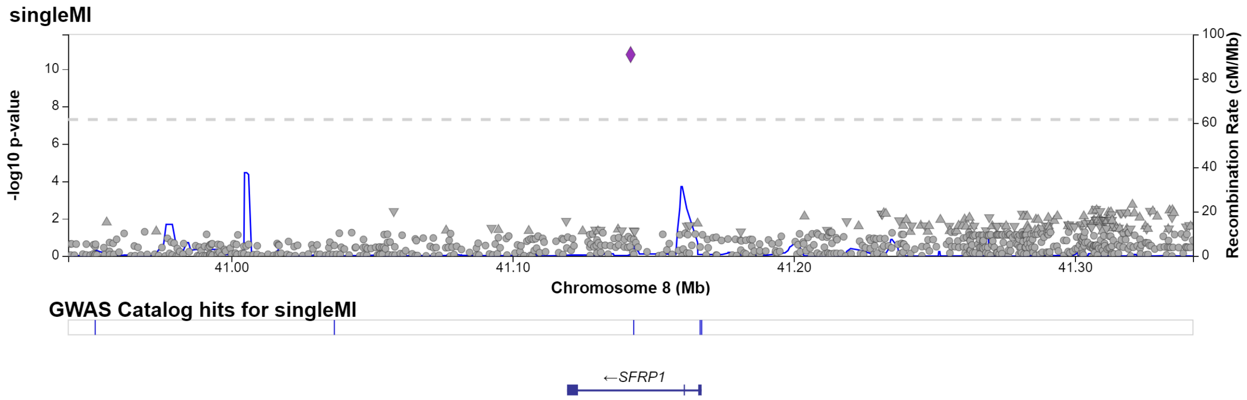


Fig. S30: Regional plot of single significant hit at chromosome 8 for single MI.

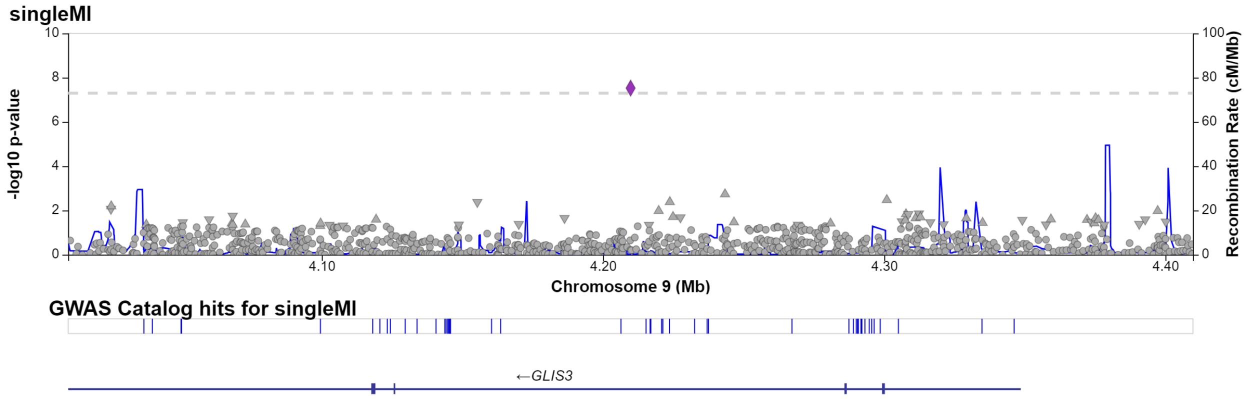


Fig. S31: Regional plot of single significant hit at chromosome 9 for single MI.

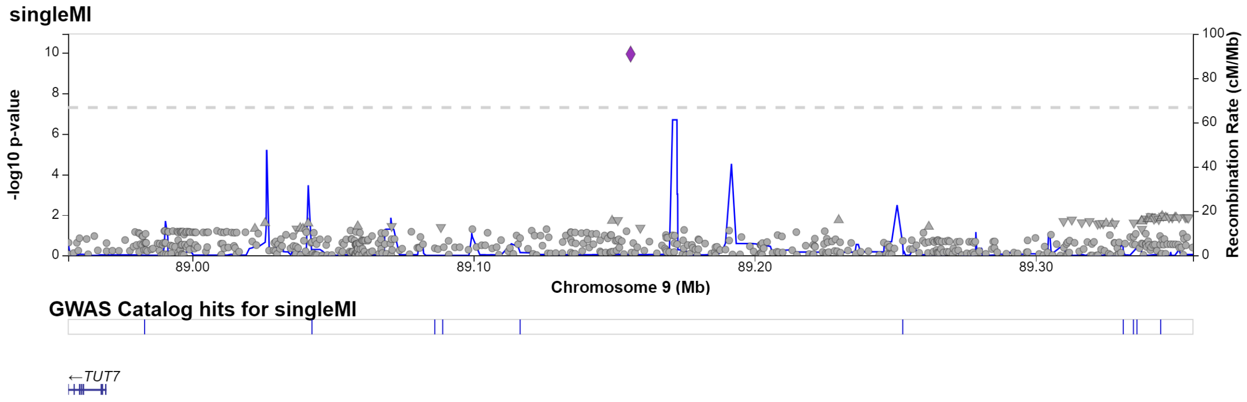


Fig. S32: Regional plot of single significant hit at chromosome 9 for single MI.

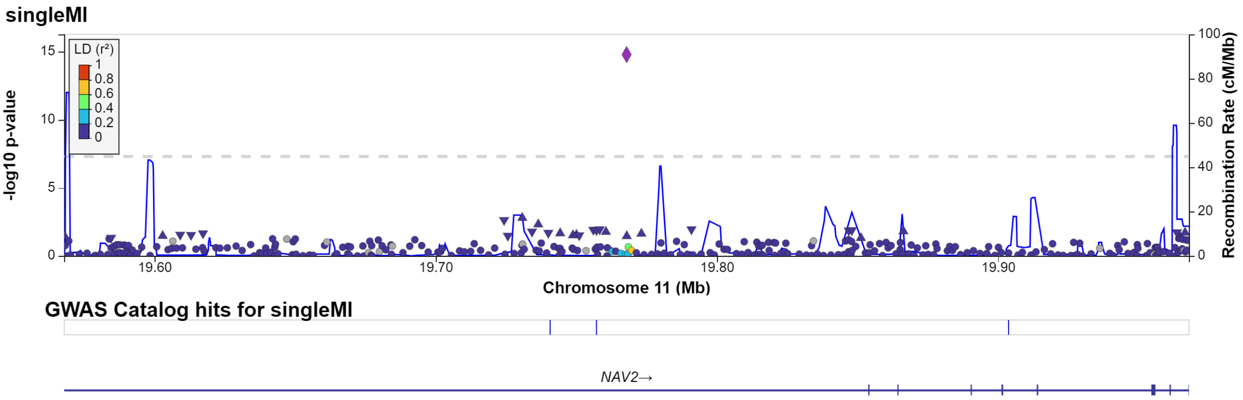


Fig. S33: Regional plot of single significant hit at chromosome 11 for single MI.

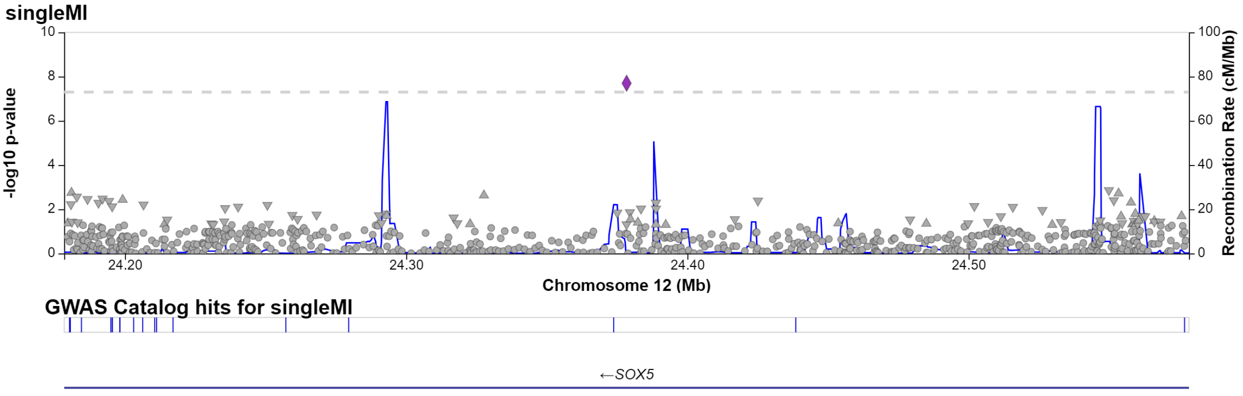


Fig. S34: Regional plot of single significant hit at chromosome 12 for single MI.

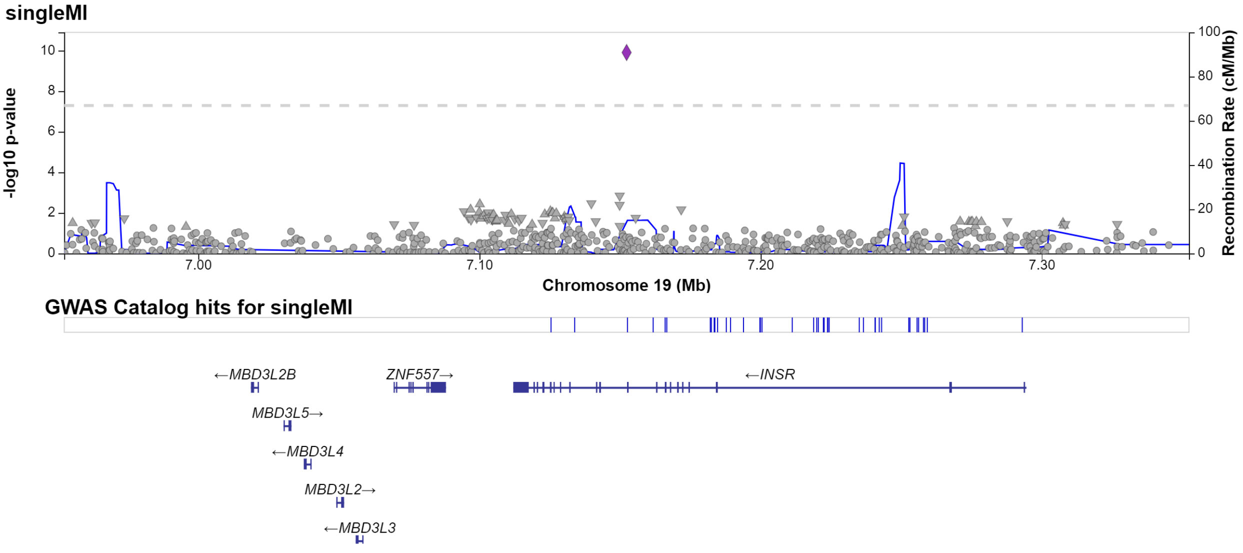


Fig. S35: Regional plot of single significant hit at chromosome 19 for single MI.

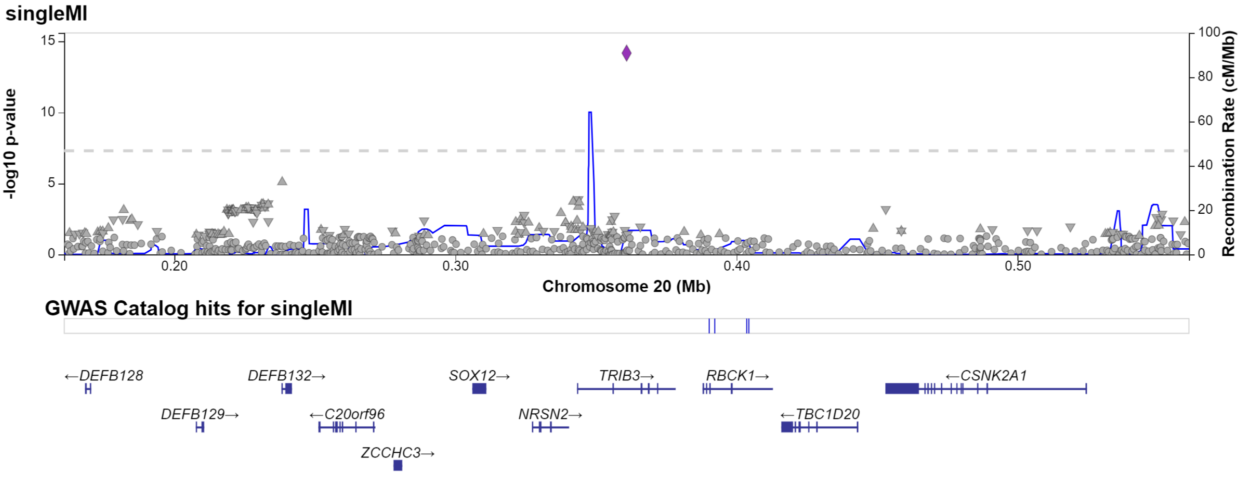


Fig. S36: Regional plot of single significant hit at chromosome 20 for single MI.

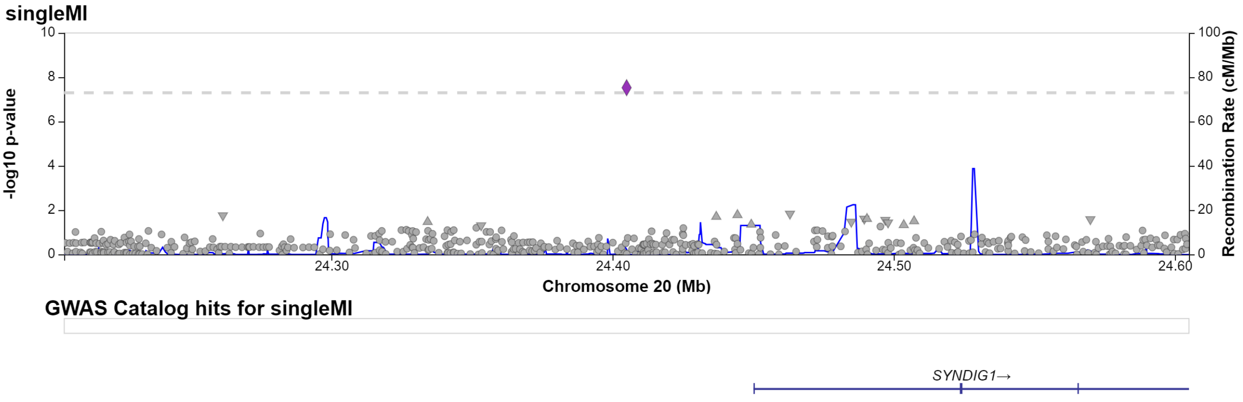


Fig. S37: Regional plot of single significant hit at chromosome 20 for single MI.

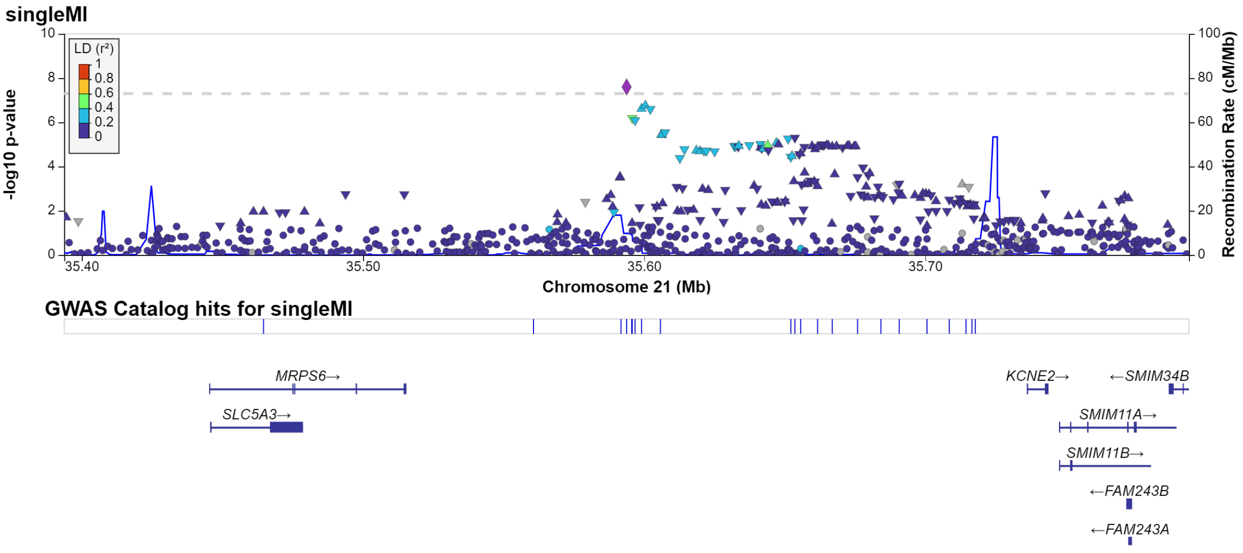


Fig. S38: Regional plot of single significant hit at chromosome 21 for single MI.

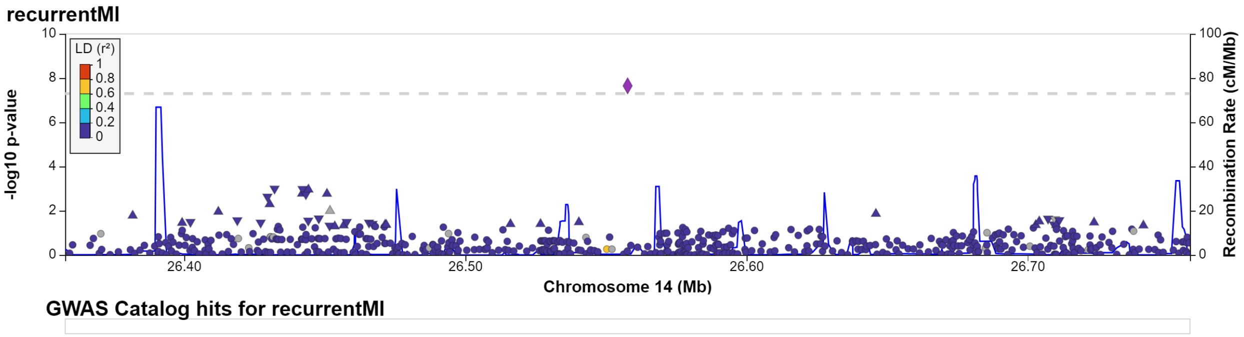


Fig. S39: Regional plot of single significant hit at chromosome 14 for recurrent MI.
